# Scale-free dynamics of Covid-19 in a Brazilian city

**DOI:** 10.1101/2021.09.10.21263332

**Authors:** J. M. P. Policarpo, A. A. G. F. Ramos, C. Dye, N. R. Faria, F. E. Leal, O. J. S. Moraes, K. V. Parag, P. S. Peixoto, E. C. Sabino, V. H. Nascimento, A. Deppman

## Abstract

Mathematical models can provide insights into the control of pandemic COVID-19, which remains a global priority. The dynamics of directly-transmitted infectious diseases, such as COVID-19, are usually described by compartmental models where individuals are classified as susceptible, infected and removed. These SIR models typically assume homogenous transmission of infection, even in large populations, a simplification that is convenient but inconsistent with observations. Here we use original data on the dynamics of COVID-19 spread in a Brazilian city to investigate the structure of the transmission network. We find that transmission can be described by a network in which each infectious individual has a small number of susceptible contacts, of the order of 2-5, which is independent of total population size. Compared with standard models of homogenous mixing, this scale-free, fractal infection process gives a better description of COVID-19 dynamics through time. In addition, the contact process explains the geographically localized clusters of disease seen in this Brazilian city. Our scale-free model can help refine criteria for physical and social distancing in order to more effectively mitigate the spread of COVID-19. We propose that scale-free COVID-19 dynamics could be a widespread phenomenon, a topic for further investigation.

---

The pandemic of SARS-COV-2 has had a huge impact on our way of life (1, 2). In many countries, social distancing, lockdowns or quarantines were adopted to diminish the rate of new cases of infection. These decisions were, in many cases, successful in controlling the fast increase in infection and in avoiding a potential collapse of health systems (3–6).

The usual approach to modeling of epidemic spread is with compartmental models such as the Susceptible-Infected-Recovered (SIR) model or variations thereof (7–11). A simpler formulation based on similar assumptions is the Susceptible-Infected (SI) model (12). The basic assumption is that one infected (I) individual can transmit, with a given probability, the virus to any other susceptible (S) individual in the population. The consequence of this assumption in these models is an exponential growth in the number of infected individuals at the beginning of an outbreak. However, analyses of previous epidemics (13), and of COVID-19 in particular, show that the early growth of infected cases tends to be subexponential, following a power-law (14–18).

Complex Networks, where agents are the nodes of the network, and the contacts among agents are represented by edges, have been studied in detail in recent years (19) and might also be useful to describe the spread of infection. The effects of agents at different scales were observed by the identification of household bubbles in the epidemic process in the case of the COVID-19 epidemics (20). The special case of scale-free networks has received attention due to the simple mechanisms that govern their growth (21). The simple assumption that new nodes will connect to the existing nodes by a preferential attachment rule generates a scale-free or scale-invariant network. The attachment rule is a non-uniform probability distribution to attach a new node to an existing node *i*. To form scale-free networks, preferential attachment must be given by a power-law distribution such that the probability of a new node attaching to an existing node *i* in a network is

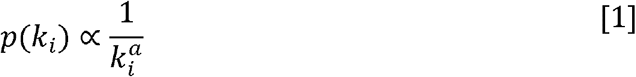

where *k*_*i*_ is the number of edges connecting the *i*th node and *a* is a constant. Several aspects of scale-free networks follow power-laws, and scale-free networks can be obtained under a variety of attachment rules (22).

Scale-free networks obtained by preferential attachment mechanisms have the property of self-similarity (23), which is a predominant feature of fractals. Self-similarity means that a complex system, with an internal structure formed by an arbitrary number of components, is such that the components are identical to the main system, when viewed on any scale. When a network is self-similar, cutting any branch of the network reduces the size of the network but leaves the structure of the network unchanged.

The relations between fractal dynamics and power-law distributions were first identified by Kopelman in a study of chemical reaction dynamics (24), where the reaction rate followed a power-law dependence on time. Complex networks are useful tools to investigate epidemic processes too (25–27), and the fractal dynamics approach has already been used to study Covid-19, where the fractal phenomenon was expressed by a power-law in the time dependence of one of the parameters of a SIR model. The model proposed in (14) follows a different approach. It assumes fractal dynamics to extend the mechanisms of infection in small groups to larger groups, using the self-similarity typical of fractals. In the present work, we show that such an assumption will lead to a power-law distribution of the parameters of the model.

Roughly speaking, the fractal approach assumes scale invariance of the complex structures of the epidemic dynamics, with different groups of infection in a given population. With such an assumption, the dynamics of the epidemic spreading over a large population can be related to that of a number of small contact groups, and the number of parameters necessary to describe the complex dynamics is reduced, simplifying the task of model fitting. In the model used here, “agents” designate the groups of close contacts into which the population is divided. Each agent is formed by other agents at smaller scales, until one reaches the smallest agent, that is a group of individuals who have the most frequent and close contact. The fractal aspect is introduced by considering that any relevant property of the epidemic process in a large agent can be described by the same dynamic that happens in a smaller agent, after the quantities are scaled to the smaller population. In other words, the agents are self-similar.

With the self-similar assumption, the number of parameters of the model is reduced, and the modes of contagion in a large population can be investigated by observing what happens in a small group. By “modes of contagion” we mean the different ways in which an initially infected agent may infect a second agent in the same contact group (larger agent): Considering the group of four agents in Figure (1), the infectious agent (red) can infect the agent on the left (a) directly, as shown in the left; (b) indirectly, through a third agent, as shown in the middle; (c) indirectly, through two other agents, as shown in the right. Note that there are three possible chains of direct infection: C-L, C-B, C-R, three chains of secondary infection: C-L-R, C-L-B, C-B-R (considering only the subgroups of infected agents, and disregarding the order of infection), and only one chain of third-order infection C-L-B-R. We call these possibilities “modes of infection” If *τ* is the fraction of infected agents in the group per contact, and assuming that *τ* ≪ 1, it can be shown that the number of infected agents, *v*, during the period of infection in the group is given by (14)

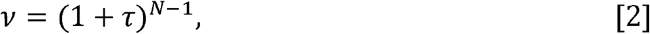

where *N* is the number of agents in the group.

**Fig. 1.**
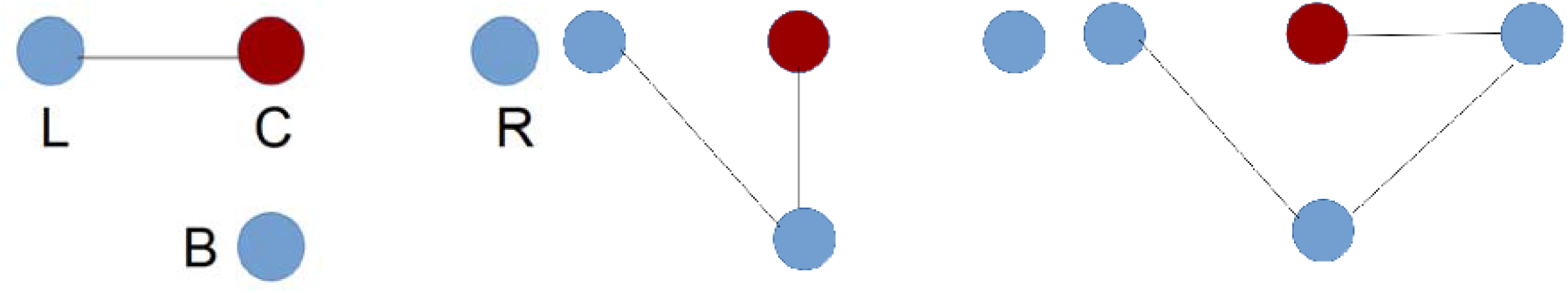
Modes of contagion in a small group of four individuals ((L)eft, (C)enter, (R)ight and (B)ottom). The figures show schematically the different modes, direct (left panel) or indirect (central and right panels), that one susceptible individual can be infected in the group if one individual is initially infected.

The agents in the smallest groups have size *λ*_*o*_ = 1, i.e., they correspond to individuals. Equation (2) describes disease transmission inside these smallest group of individuals. The scaling is made by assuming that a larger group, with population *u*_*λ*_, is formed by *N*_*λ*_ agents of size of the order of *λ*, so that *u*_*λ*_ ∼ *N*_*λ*_ *λ*, and modifying Equation (2222) to

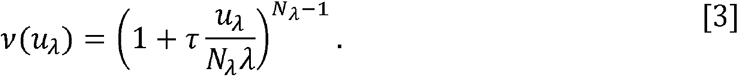

The *q*-exponential is usually written in the form

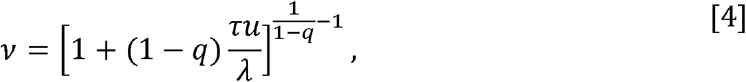

where 1 − *q* = 1/*N*. In the epidemic process, this is the contribution of one single agent, so the total number of infected individuals is obtained by integrating the expression above over the entire population. The number of infected agents, therefore, is of the form

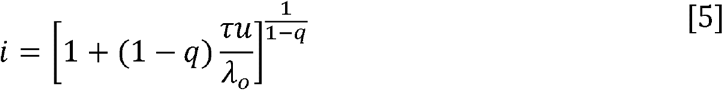

In this fractal approach to the epidemic dynamics, it is assumed that the average number of contacts made by an individual is not strongly dependent on the population of the region in which this individual is living, that is, if they live in a large metropolis with around 10^7^ inhabitants, or if she lives in a small city with 10^3^ inhabitants, her number of contacts will always be around a few tens of individuals. The meaning of close contacts may vary depending on the aspect of social life one is interested in, and since here we analyse virus spread, by close contact we mean those individuals with a frequent and in-person contact, with possibility of virus transmission. In this work, we initially consider an average number of contacts for the whole population (i.e., constant *q*_*j*_), but later we investigate how small variations, relative to the total population, of the number of contacts of different agents can be observed, and we will identify some super-spreader agents.

The q-exponential function in Equation (5) is typical of the non-extensive statistics developed by Tsallis (29), a generalization of the Boltzmann-Gibbs statistics that has found applications in many fields, including fractals (30). The fact that it appears here is an indication of the adequacy of the non-extensive statistics to describe the epidemic dynamics. The relations between fractal structures and Tsallis statistics have been investigated in other works (30, 31).

So far, we have said nothing about the evolution of the epidemic through time. In the model proposed in Ref. (14), time dependence is introduced by assuming that the infection in a single agent *j* evolves by homogeneous mixing over a time *Δt*_*j*_ =*t*_*fj*_ − *t*_*oj*_, where *t*_*oj*_ and *t*_*fj*_ are, respectively, the time of start of the epidemic and end of the epidemic process in that agent, the last one corresponding to the instant when 95% of the agent population is infected or removed.

Within the assumptions of the model up to this point, *u*_*j*_ represents the number of agents that are in close contact in a group with an infected agent. To allow part of the population to remain without being infected, two modifications are added to the model: the population in a group, *s*_*j*_, can be larger than *u*_*j*_, and a new parameter, *κ*\*, regulates the fraction of the total population that evades infection at the end of the process. Define the rate of contagion of the agent, *κ*_*j*_, such that

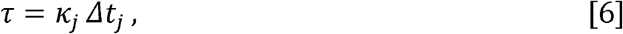

and obtain the time-dependent function of the number of infected individuals in agent *j*,

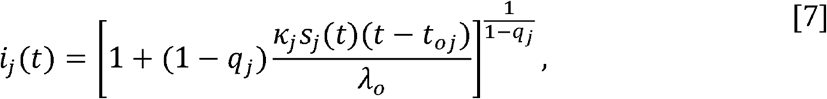

where the susceptible population is considered to be a function of time, *u*_*j*_(*t*), since those individuals that were infected are not considered susceptible anymore.

The model can be described by the coupled differential equations (see the Appendix for details)

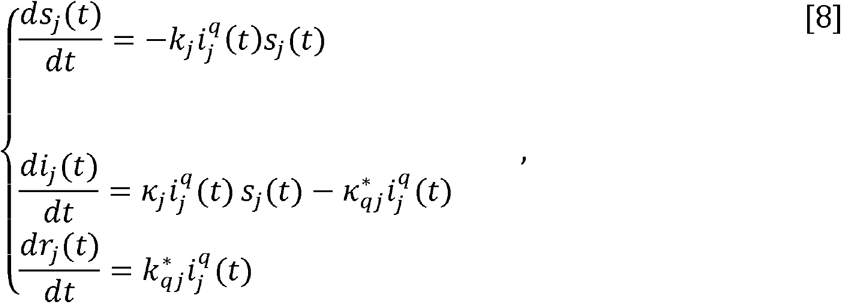

for *t* ≥*t*_*oj*_. Here, *s*_*j*_, *i*_*j*_ and *r*_*j*_ represent, respectively, the susceptible, infectious and recovered populations of agent *j*. The parameters *κ*_*j*_ and 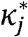 are the rate of infection and the rate of removal, respectively. Observe that the rate of infection can appear in a normalized form, as *κ*_*j*_ = *β*_*j*_/*s*_*jo*_, where *s*_*oj*_ is the total population of the agent *j*. The derivation of the set of equations above from the basic principles of the model is developed in the Appendix. The rate of removal is given by

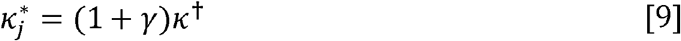

With

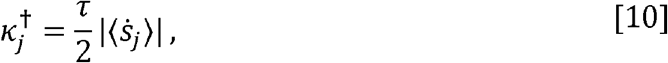

where 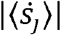 is the average rate of new infections during the course of the epidemic in a particular agent (see Appendix). Note that 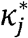 is completely determined by the other parameters of the epidemic dynamics, with *τ* = *κ*_*j*_ *Δt*_*j*_. The parameter *γ* allows the inclusion of additional mechanisms in the spreading process, as the removal of individuals before all other individuals in the agent are infected, and is determined on basis of additional information, as the total population of the agent.

Equations (8) are similar to the SIR equations, aside from the exponent *q* in the infected population *i*_*j*_(*t*). A phenomenological approach introduced a similar set of equations with powers on the population of infected individuals (32), but here those equations result from a clear assumption on the fractal dynamics of the epidemic process. The exponent *q* is a fundamental aspect of the Tsallis statistics.

There are two forms for the increase of the number of infected individuals within an agent: by the time evolution of the spreading, with a rate that is described by the derivative of *i*_*j*_(*t*); or by merging with another agent when their individuals cannot be clearly separated. In this case, the mathematical description is done by using so-called q-algebra, which is associated with Tsallis statistics, and which keeps a fixed value for q while summing the populations of the merging agents.

The infected population, as a function of time, is

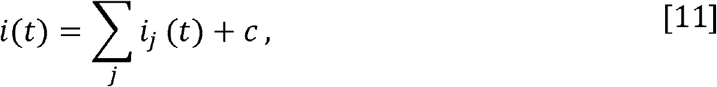

where *c* is a constant representing the average daily number of new cases that come from outside and seed new clusters of infection in the city.

Here there is an important difference between the fractal model and the traditional models: rather than an exponential increase in the number of new cases, a slower increase, following the q-exponential function, is found whenever *q* ≠ 1. Only in the case of *q* = 1 the SIR and SI equations are recovered.

## Results and discussion

We now fit the fractal model Equation (8) to detailed data from the COVID-19 epidemic in the city of São Caetano do Sul, Brazil, in which residents with suspected COVID-19 symptoms were encouraged to contact a dedicated group that offered a home visit for a PCR test (see Section supplement). We perform an analysis in four parts:

1. We estimate the number of agents and the parameters *u*_*j*_, *κ*_*j*_ and *Δt*_*j*_, assuming the fractal model with 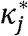 determined by the other parameters, following Equations (9)–(10). Since previous analyses of COVID-19 showed that the number of infected individuals depends on the population as a power-law with exponent 1.15 (14), we use *q* = 0.13, which results in an exponent 1/(1 − *q*) =1.15 in Equation (7). This exponent of the power-law is compatible with the observations of several quantities related to life in cities (33–35). The goal of the analysis here is to determine the rates with which parameters *κ* and *Δt* change as the population grows, which allows us to evaluate the dynamics of the epidemic and estimate the value of *q* in Section supplement (see also the discussion at the end of Section supplement). The analysis uses four different choices of time resolution, gathering the data in bins of widths 4, 7, 15 and 30 days. When the number of new cases is grouped into larger bins, the data is associated with larger groups of infection, and fewer details of the evolution of the epidemic can be observed in the data set. In the context of fractals, the use of different bin sizes is associated with a change in the scale of the system, so larger bins correspond to larger groups of infection. This results in models in which the number of susceptibles is approximately equal to the number of infected observed in the data (which is *N*_infected_ = 2,573).
2. Having estimated the parameter *κ*_*j*_ for larger agents in Part 1, we now fit a different value of *q*_*j*_ for each agent in each scale. We obtain an estimate for the social distancing, and compare our results to an independent social isolation index obtained from cellular phone data.
3. In this part we use the geographical information about approximate residence location of each individual in the data set to create clusters of infection in space and time.
4. In the last part of the analysis we compare the ability of the fractal model and the SIR model (*q* → 1) to fit the data.

### Results for Part 1

The analysis here, as described in the Supporting Information Appendix (SIA), uses different levels of time resolution. The use of different time scales to investigate the properties of the epidemic process is not new and has been used to make predictions of the process at time *t* based in the time series up to *t* − 1 (36). Here the different scales (as mediated by the bins) are used to identify the groups throughout the outbreak that have similar properties of infection, such as the reproduction number. A lower time resolution (larger time bins) results in a smaller number of larger agents. We show in Figure (2) the best-fit results, considering 30-day and 4-day bins, with the total number of infected individuals and the contribution of each agent and the background. Table (2) presents the best-fit parameters to all the bin sizes analyzed in the present work (we use *u*_*oj*_ to denote *u*_*j*_(*t*_0*j*_)). Additional plots are shown in the SIA. We observe a good agreement between calculation and data for all cases, as evidenced by the *χ*^2^ obtained.

**Table 1.**
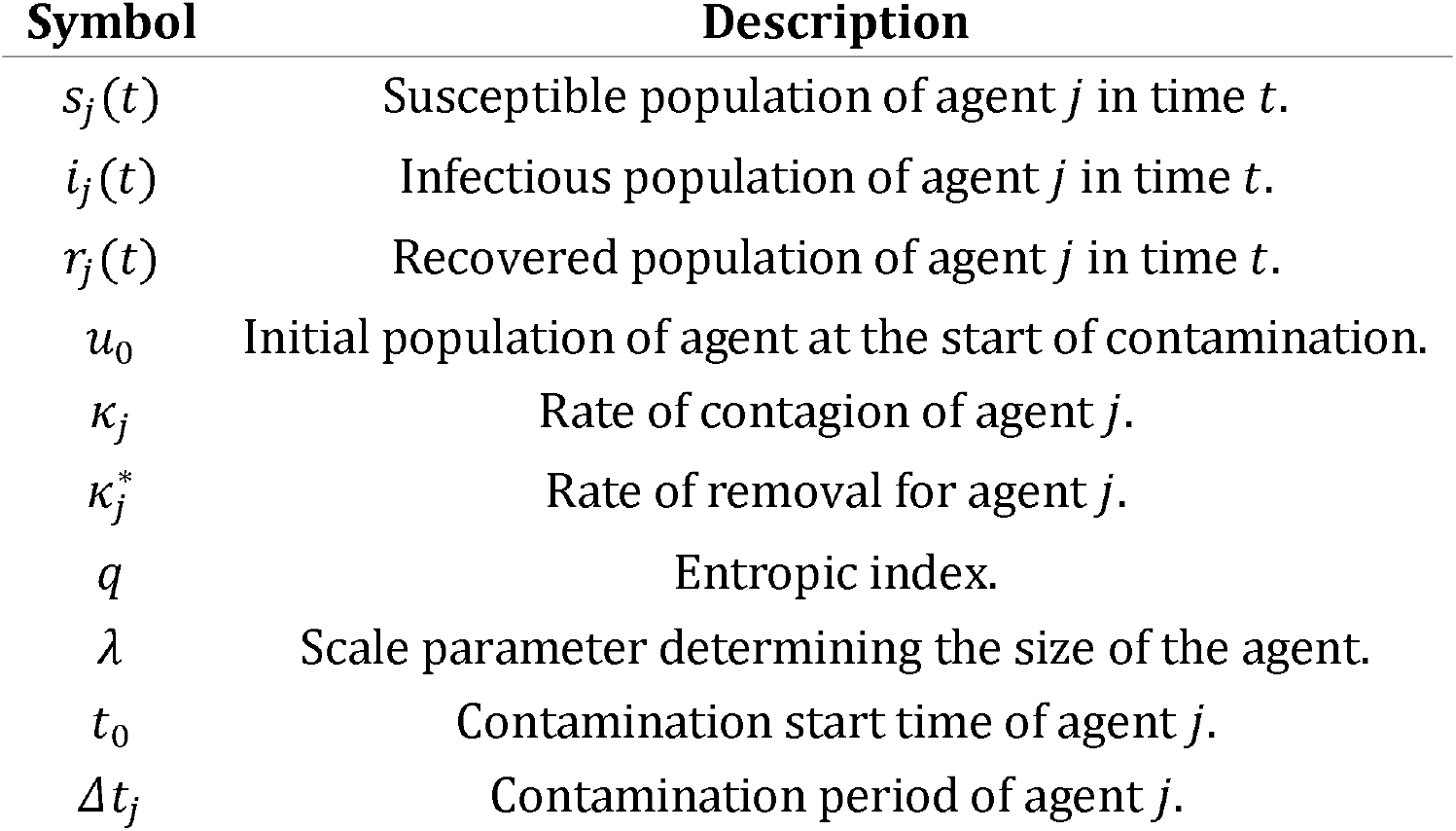
Symbol Table

| Symbol | Description |
| --- | --- |
| $s_j(t)$ | Susceptible population of agent $j$ in time $t$. |
| $i_j(t)$ | Infectious population of agent $j$ in time $t$. |
| $r_j(t)$ | Recovered population of agent $j$ in time $t$. |
| $u_0$ | Initial population of agent at the start of contamination. |
| $\kappa_j$ | Rate of contagion of agent $j$. |
| $\kappa_j^*$ | Rate of removal for agent $j$. |
| $q$ | Entropic index. |
| $\lambda$ | Scale parameter determining the size of the agent. |
| $t_0$ | Contamination start time of agent $j$. |
| $\Delta t_j$ | Contamination period of agent $j$. |

**Table 2.**
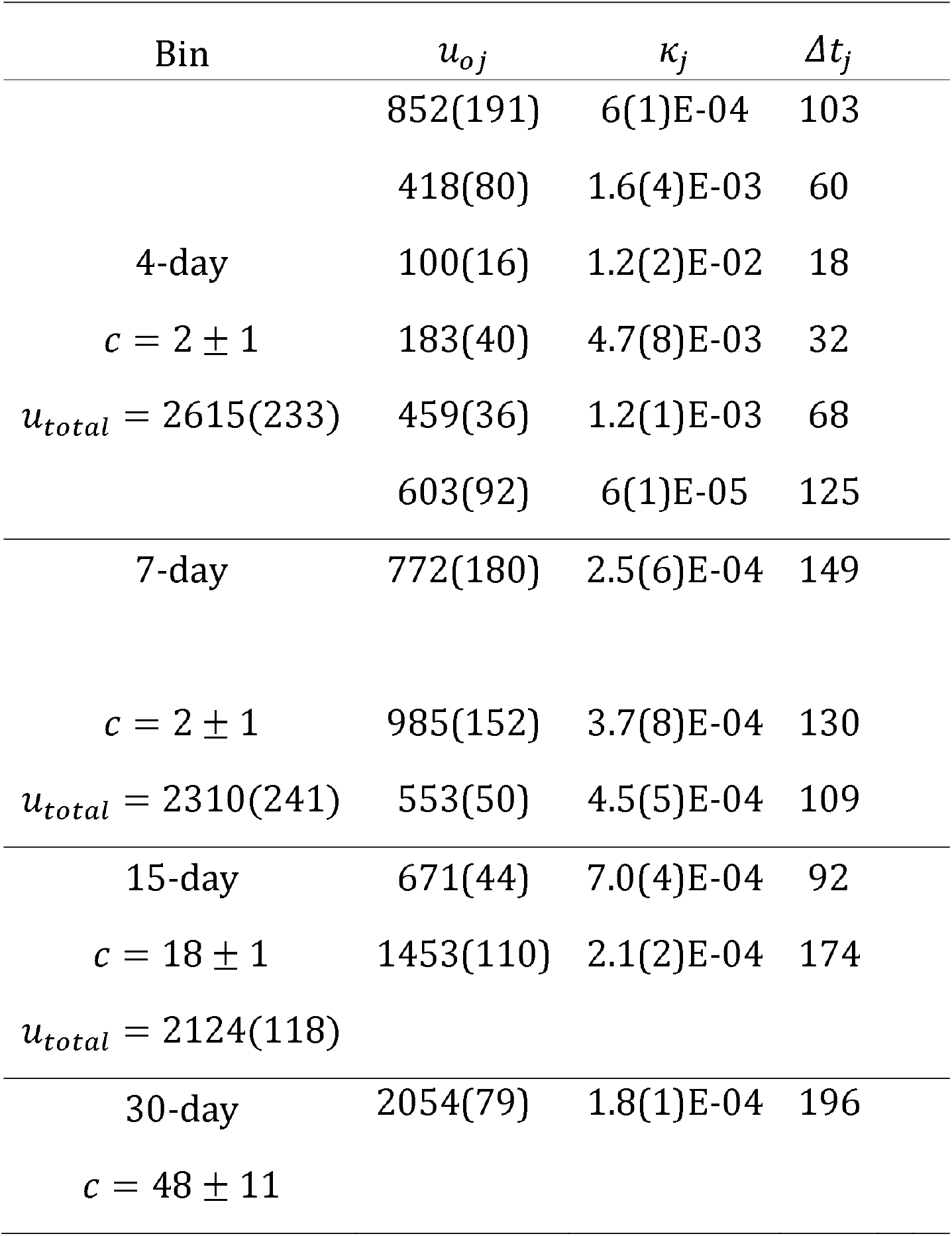
Parameters of Equation (7) adjusted for the data set separated in 4, 7, 15 and 30 day bins. The dates refer to the beginning of each group between the dates 2020-03-18 and 2020-09-19 as shown in Figure (2). The background **c** is a constant adjusted to the whole period in each case.

**Fig. 2.**
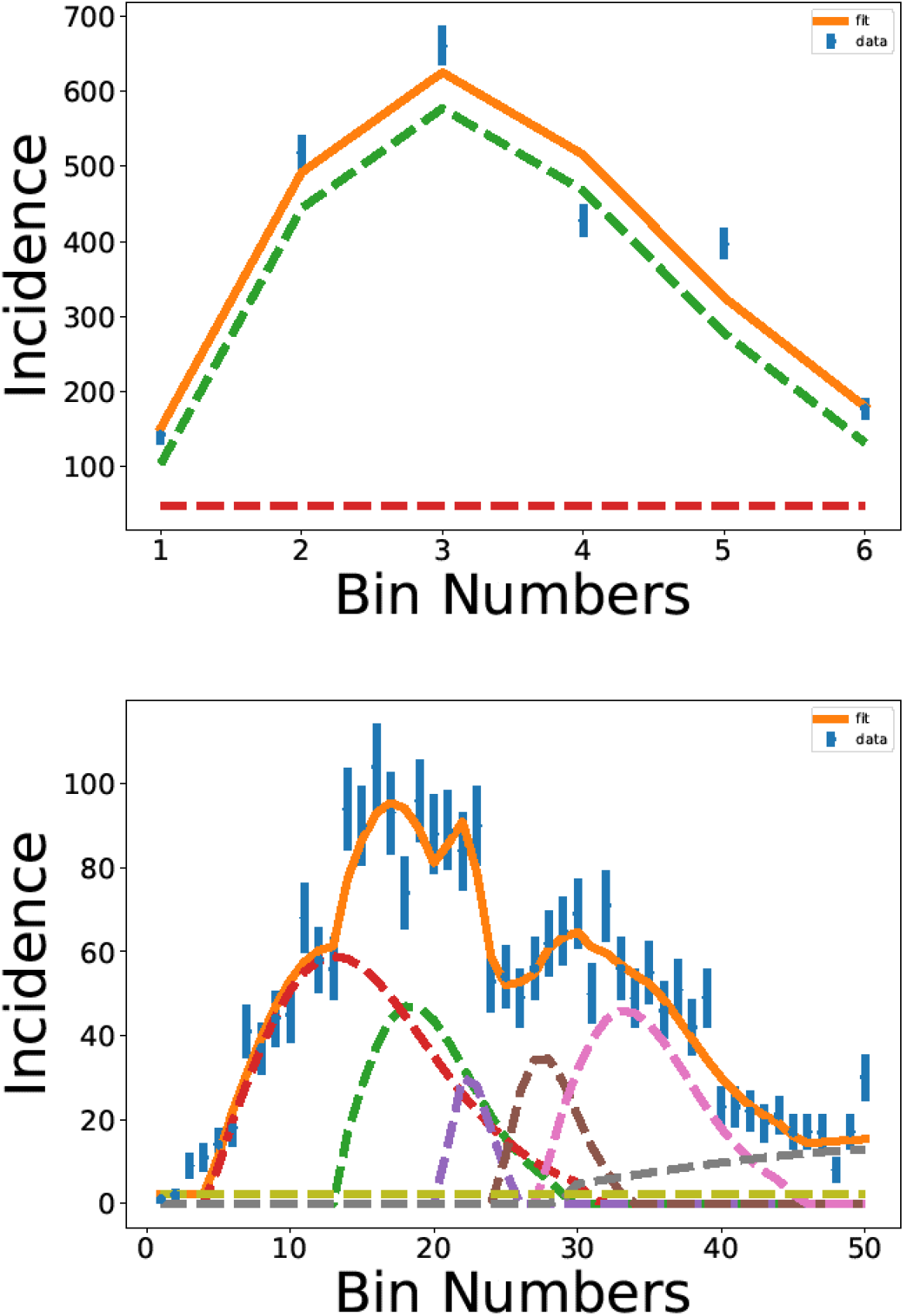
The results of the fitting procedure for (top) 30-day bin and (bottom) 4-day bin analysis. The total number of infected (continuous line) and the contributions of each agent (dashed lines) are shown, as well as the background contribution indicated by the horizontal line. The parameters for each peak are shown in ordered by the initial time of spreading in each agent.

The complexity shown by the data increases as the bin width is reduced, and for the 4-day bin analysis, the complex pattern of fluctuations represents a challenge for the fitting procedure, with several agents needed to obtain a good description.

Nevertheless, the model successfully describes the data with six agents and a background contribution, which corresponds to the rate of people that are infected out of the city, and seeds a new group of infection in the city. At the end of this part of the analysis, we obtained a set of 12 values for the agent size, transmission rate and infection period for each agent.

With the data for the infection period, *Δt*_*j*_, *κ*_*j*_ and *u*_*oj*_ for each agent, we can analyse the behaviour of those parameters and verify if the model is consistent. In fact, as discussed in the Introduction, the variables *Δt*_*j*_ and *κ*_*j*_ must follow a power-law distribution due to the fractal dynamics, with 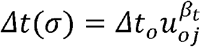 and 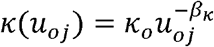. The plots in show Figure (3) the data distributed along a straight line in the log-log plot, confirming that the results obtained are consistent with the assumption made in the model.

**Fig. 3.**
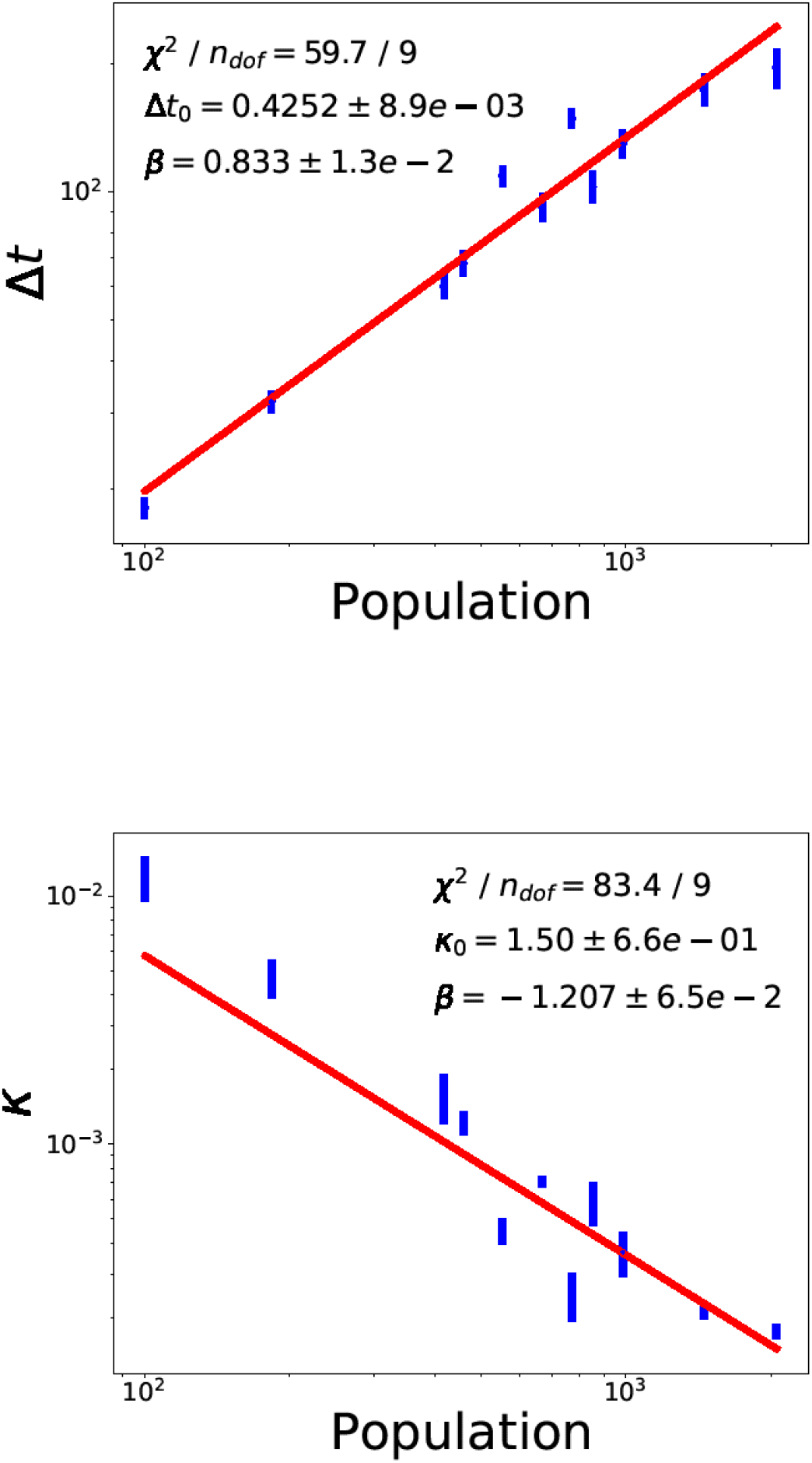
Plots of the parameters Δt and κ as a function of the agent population, as obtained in the fitting procedure.

With the results obtained for *κ* and *Δt*, the parameter *τ* = *κΔt* depends on the population size, although it was initially supposed to be constant. This dependence of *τ* on the population size indicates that the value chosen for *q* is not correct for describing COVID-19 evolution in the city studied here. In the next section we show that the value for *q* is different from the value adopted in this part of the analysis. To obtain this result, we will use the fact that *κ* must behave as a power-law, and we adopt the exponent *β* = −0.833, which keeps the value of *τ* constant.

### Results for Part 2

In the second part of the present study, the power-law for 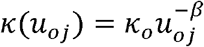 is adopted with *β* = −0.833, while the parameter *q* is free to adjust the data. We also analysed the data with *β* = −1.2 and found that the main conclusions would be the same as obtained in the case adopted here. For instance, the fact that the social distancing observed, as described below, will decrease between the first and the second wave, stands with either value for *β*. Observe that we expect *τ* to be approximately independent of the agent considered, since this parameter should capture the characteristics of the virus, while *q* is supposed to reflect the characteristics of social behaviour. As discussed above, virus characteristics and social behaviour cannot be completely separated, so variations on the value for *τ* may be expected. Therefore, we do not expect a well defined behaviour of the parameter with the agent size, as would result from choosing *β* = −1.2 for *κ*_*oj*_, which would lead to 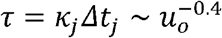. We attribute this result to the value chosen for *q*, which might be slightly different of the value corresponding to the average number of contacts the population has, as we investigate below. Therefore, we adopt for *κ* a behavior that corresponds to the inverse of that observed for *Δt*, and fix its exponent to the value mentioned. We choose this value because the interval *Δt*_*j*_ results from the good fitting to the data, and is determined with better precision that the values for *κ*_*j*_. This aspect is discussed in more details in the SIA. The results of the fitting procedure are shown in Figure (4a), where we see the first wave, used in the previous analysis, and the peaks corresponding to the second wave of infection in the city. We notice that the second wave presents peaks that are narrower than those in the first wave, indicating that the process, in this case, is somewhat different from the previous wave. The best-fit parameters are presented in Table (3). Additional plots are available in the SIA.

**Table 3.** Best-fit parameters of Equation (7) in first wave obtained with bins of 4, 7, 15 and 30 days, and for the second wave between the dates 2020-09-19 and 2020-11-19 for bins of 4 and 7 days. The background **c** is a constant adjusted to the whole period in each case.

| global parameters | $u_{oj}$ | $q_j$ | $\Delta t_j$ | $\tau_j$ |
| --- | --- | --- | --- | --- |
| 1st wave |  |  |  |  |
| 4-day bin | 554(1) | 0.5712(4) | 82(1) | 0.0297(2) |
| $\kappa_0 = 0.06998(49)$ | 163(1) | 0.435(2) | 50(1) | 0.0501(3) |
| $c = 4 \pm 1$ | 485(1) | 0.57(1) | 78(1) | 0.0314(2) |
| $\chi^2/n_{\text{dof}} = 36/29$ | 320(1) | 0.4444(2) | 69(1) | 0.0395(3) |
| $u_{\text{total}} = 2084(2)$ | 652(1) | 0.5388(3) | 89(1) | 0.0282(2) |
| 7-day bin | 569(35) | 0.19(1) | 76(3) | 0.0398(9) |
| $\kappa_0 = 0.08723(43)$ | 1170(1) | 0.2850(4) | 90(1) | 0.0281(3) |
| $c = 9 \pm 2$ | 402(27) | 0.44(7) | 72(1) | 0.0401(1) |
| $\chi^2/n_{\text{dof}} = 35/15$ | | | | |
| $u_{\text{total}} = 2141(44)$ | | | | |
| 15-day bin |  |  |  |  |
| $\kappa_0 = 0.07(1)$ | | | | |
| $c = 23 \pm 1$ | 1701(355) | 0.42(4) | 134(1) | 0.020(4) |
| $\chi^2/n_{\text{dof}} = 13/4$ | 342(150) | 0.5(2) | 67(1) | 0.04(1) |
| $u_{\text{total}} = 2043(385)$ | | | | |
| 30-day bin | 2255(173) | 0.6(2) | 190(1) | 0.012(6) |
| $\kappa_0 = 0.04(2)$ | | | | |
| $c = 33 \pm 29$ | | | | |
| $\chi^2/n_{\text{dof}} = 34/1$ | | | | |
| 2nd wave |  |  |  |  |
| 4-day bin |  |  |  |  |
| $\kappa_0 = 0.48071(15)$ | 114(11) | 0.429(4) | 15(1) | 0.14(1) |
| $c = 13 \pm 1$ | 67(1) | 0.756(8) | 9(1) | 0.1370(7) |
| $\chi^2/n_{\text{dof}} = 14/3$ | 376(1) | 0.472(1) | 24(1) | 0.0819(2) |
| $u_{\text{total}} = 557(11)$ | | | | |
| 7-day bin |  |  |  |  |
| $\kappa_0 = 0.172520(13)$ | 217(2) | 0.602(1) | 33(1) | 0.0645(9) |
| $c = 23 \pm 1$ | 408(1) | 0.9647(7) | 34(1) | 0.03922(4) |
| $\chi^2/n_{\text{dof}} = 6/1$ | | | | |
| $u_{\text{total}} = 625(2)$ | | | | |

**Fig. 4.**
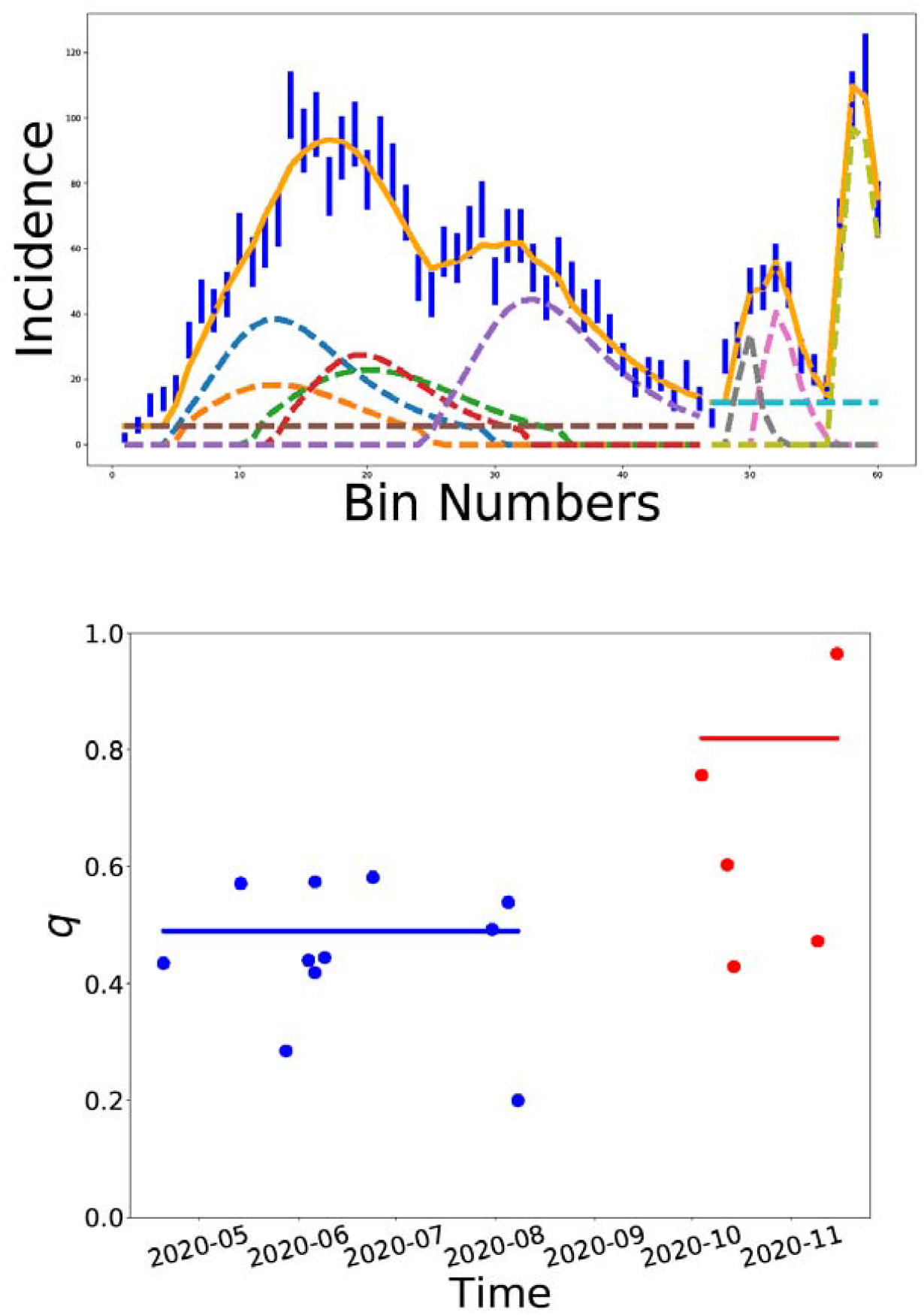
The data grouped in bins of 4 days, with q free to adjust the data and using the power-law behaviour for κ in the first and second waves between 2020-03-18 and 2020-09-19 (top). The value for q as a function of time using the power-law κ = κ_0_u^−0.8.3.3^ (bottom). In blue are the parameters found in the first wave and in red for the second wave.

In the first part of the analysis, we checked the consistency of the fractal hypothesis by verifying that the parameters *κ* and, *Δt*, vary with the agent population according to power-laws. After finding that the hypothesis of fractal dynamics is consistent with data we now allow *q*_*j*_ to be a free variable adjusted to fit the data. We observe that also in this case the model can describe the data in detail. The parameters *q*_*j*_ and *u*_*oj*_ are plotted in Figure (4b), where we can observe the data of the first wave (blue online) and of the second wave (red online). The data for the first wave are concentrated in the range 0 < *q*_*j*_ < 0.6 The second wave, however, has values that are above *q* = 0.6. The parameter *q* is related to the number of close contacts of the agents in each group, given by *N* =(1 − *q*)^−1^ .So these results show that our model is capable of evaluating the social distancing based exclusively on the data of the number of infected individuals. For the first wave, we obtain *N*∼2, while for the second wave we obtain *N*∼5, indicating that the population in São Caetano relaxed social distancing during the second wave. A clear consequence is a fast increase observed in the number of infected in the second wave (Figure 4).

We observe that *χ*^2^ decreases as the number of agents increases showing, as expected, that the model provides a better description of epidemic dynamics when there are assumed to be more agents. The fact that *q* increases from the first to the second wave shows a change in the social behaviour of the population. In each period, there are groups with small values for *q*, indicating small social contact, and others with higher values for *q*. In the second wave, as we observe in Figure (4), that some agents present very high number of contacts, and can be identified as super-spreaders.

The value of *τ* varies very little for all agents in the first wave (see Figure S5 in the SIA), with ⟨*τ*⟩ = 0.03625(7). For the second wave, the value for *τ* is not as uniform, showing that the fractal model is not able to completely disentangle the properties of the virus infection contained in *τ*, from the social behaviour, contained in *q*. This difficulty of the model to separate the effects of social behaviour on the variable *τ* may be due to the fast change in the behaviour of the population in that period, which might be fast even compared to the infectious period, *Δt*, of the agents found for the 4-day bins. Another possibility is the appearance of new variants around the time of the second wave — for example, the P.2 variant seems to have appeared around mid-2020 (37), and started being reported in São Paulo state around Sept. 2020 (38).

One of the ways of verifying the intensity of the interactions between the first and second waves is by observing the degree of social isolation throughout the year and comparing it with the results obtained numerically.

In Figure (5) we show how the Social Isolation Index, indicating the percentage of people that stayed home during the respective day, varied during 2020 in the city of São Caetano. Both used datasets qualitatively agree with respect to the mobility pattern of the population. Social isolation during the first wave of the pandemic was significantly higher in the city compared to the beginning of the second wave.

**Fig. 5.**
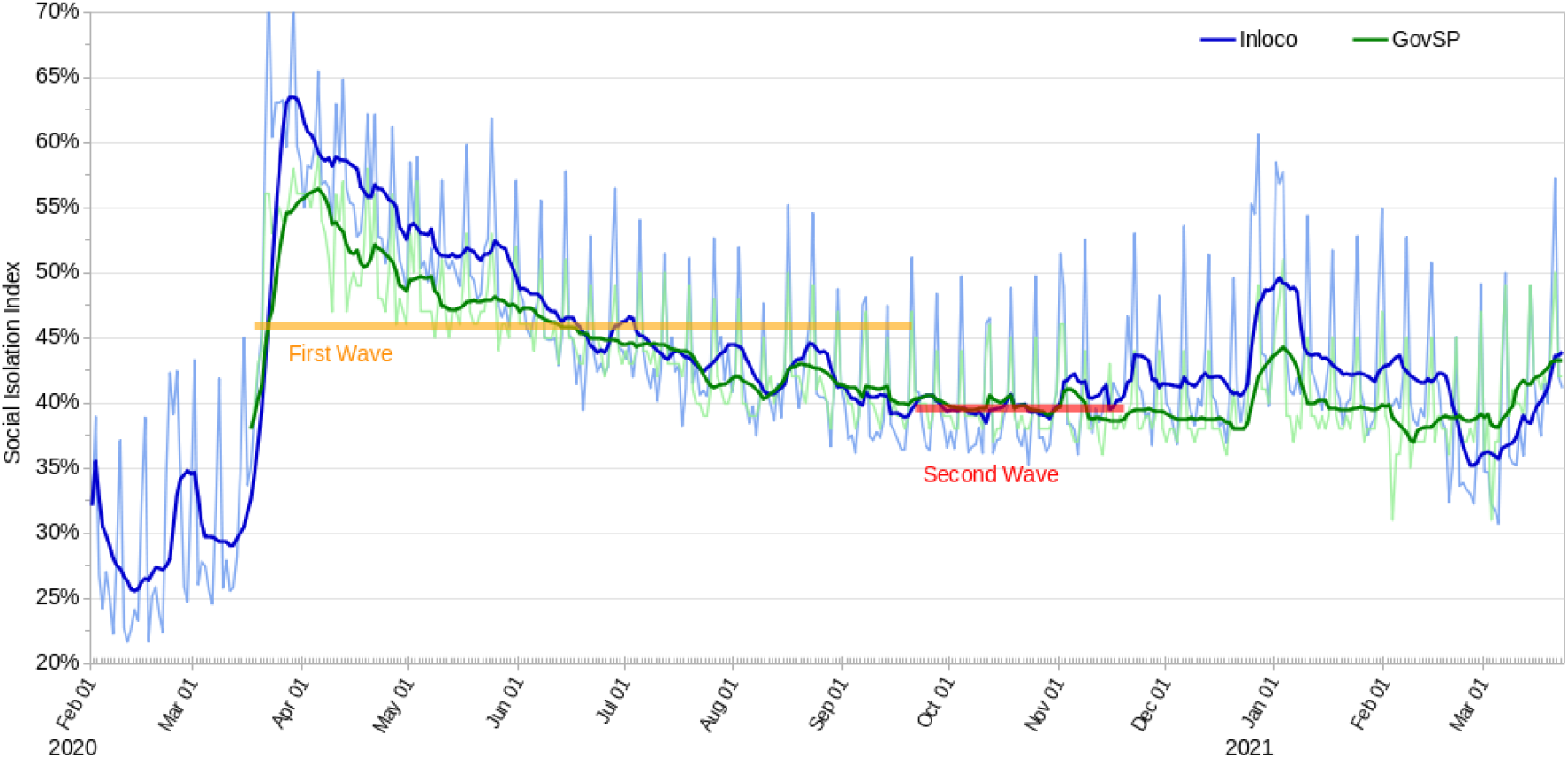
Social Isolation Metrics for the city of São Caetano do Sul indicating the percentage of people that stayed home during the respective day. Two indexes are shown: in blue, the index from the Inloco company and in green the official index from the state government of São Paulo. The daily index is shown in light colours and the seven-day moving average is shown in dark colours, for each source. Horizontal bars indicate the average social isolation index, using the official government dataset, of the first and second waves. The figure shows how adherence to social distancing policies was drastically reduced in the second wave compared to the first wave.

### Results of Part 3

The results of the third step of our analysis, where the least distance between newly infected individuals and the previous cases are computed, are presented in Figure (6a). We observe a concentration of cases at distances around 250 m of the previous cases even at the beginning of the infection process. This correlation in the location of residence of the infected individuals is an indication of the existence of clusters of infection with geographic localization. It might be an effect of the social distancing observed at the initial phase of the first wave. The histogram for a larger period for the first wave, presented in Figure (6), shows that the short distance contagion process remains during the spreading of the disease.

**Fig. 6.**
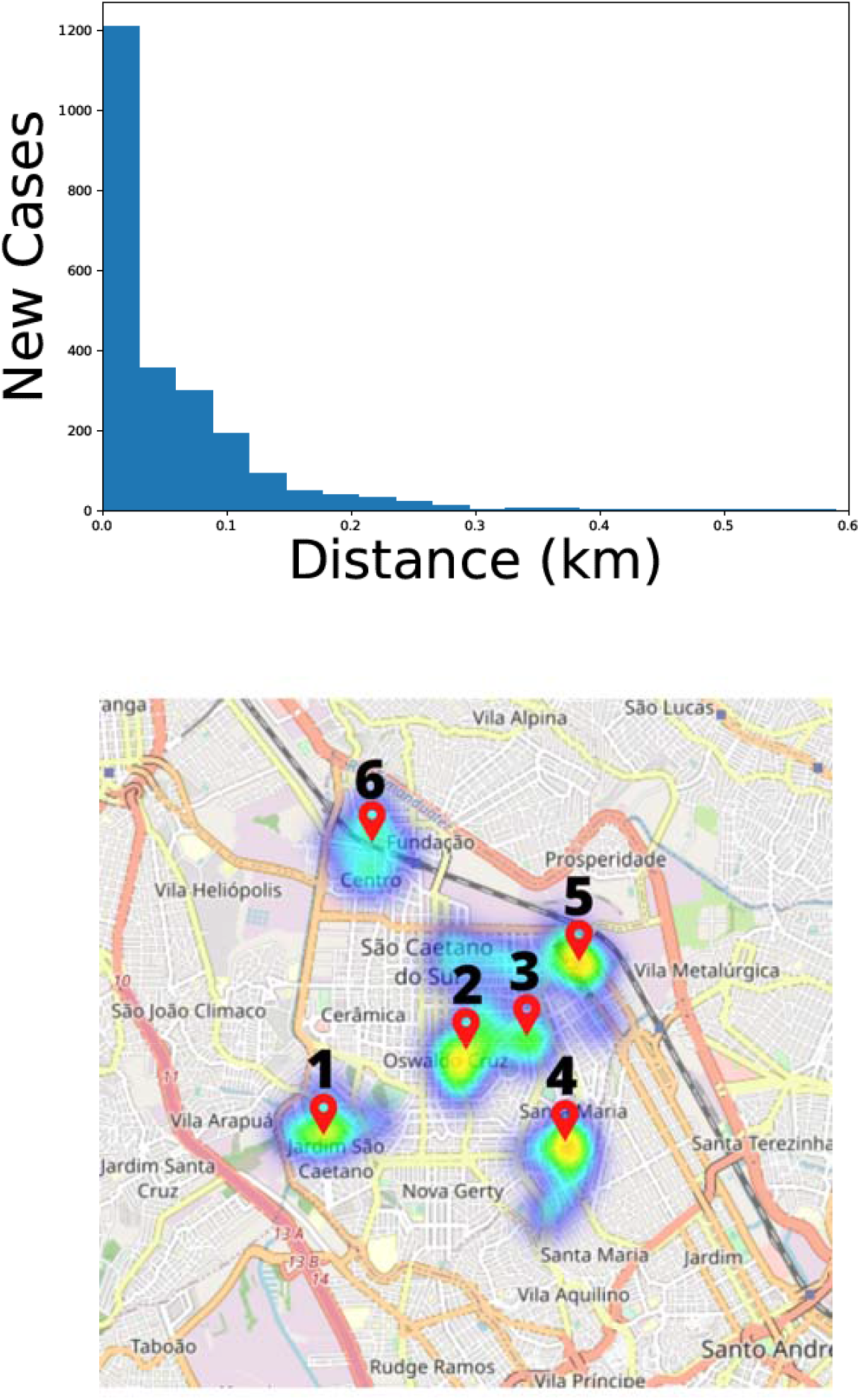
Least-distance Distribution (top) between new cases and cases from previous days. Each new case is compared with all the cases from the previous days and the shortest distance between them is chosen. The analysis was carried out over the period between 2020-03-22 and 2020-09-30 corresponding to the entire first wave. Distribution of new cases (below) using clustering method. Numbered markers represent infection groups where it is possible to associate locations of establishments with concentration of people: Technical School - near Cluster 1, Park - near Cluster 4, Municipal Stadium - near Clusters 2 and 3, Hospital - near Cluster 5, Train station and Municipal University of São Caetano - near Cluster 6, GM Factory - near Clusters 5 and 6.

With the location of residence of the infected individuals we can visualize more easily the correlations between the position of new cases and previous cases. In the SIA we present an animated-gif exhibition of the location of the new cases compared to the location of the previous cases. It is possible to observe a strong correlation between the new locations and the previous ones, reinforcing the hypothesis of close contact transmission.

Finally, with the clustering method described in the Materials and Methods section (Step 3, at the end of this document), we attribute each new case to a preexisting cluster, or to a new cluster of infection. In Figure (6b) we show the result of the clustering method for cases up to day 2020-04-20. The individuals in each cluster are presented with the same color, each color representing a different cluster. We clearly observe that the clusters are well-localized, in accordance with the least-distance findings. But in this case, the correlation is not only spatial, but takes into account the time evolution of infection. The geographic localization of the clusters across the city is shown in Figure (6b). At the beginning of the infection process, the clusters are localized, and their position in the city coincides with concentrations of people, such as universities, schools and industries. In this regard, we note that the social distancing measures adopted by the government of São Paulo started on March 23, corresponding to the initial date of our analysis, and that the results reported in Figure (6) are for the first 30 days after the date when schools and universities were closed, so the initial infections observed in the data correspond to those individuals infected before the non-pharmaceutical intervention (NPI) were put in place. Therefore, the figures show the images of the disease spreading induced by the cases of infection in the period before the closure of those institutions.

### Results of Part 4

With the equations above we can calculate the epidemic process according to different scenarios. Imposing *q* = 1 results in the SIR model, and if additionally *κ*\* = 0 we get the SI model. For *q* ≠ 1 we obtain the fractal model equivalent to SIR (*κ*\* ≠ 0) or SI (*κ*\* = 0). Below we compare and discuss the results obtained with each model.

In the following, we analyse the differences and similarities of the fractal method in comparison with the SIR model or the SI model, following the method exposed in the previous section. Initially, we fix *κ*\* = 0 and fit the derivatives of the infectious population, *di/dt*, to the data corresponding to the first wave. The result is displayed in Figure (8) and shows that a reasonable fitting is obtained with *q* = 0.56±0.03. The best-fit values for all parameters used in the different scenarios analyzed here are shown in Table (4). Observe that when all parameters are free to be adjusted, the result is compatible with *κ*\* = 0. The result of the best fit is shown in Figure (8), and can be explained by the fact leaving all parameters free results in a total population that is similar to the total number of infected individuals. This happens because the set of data, alone, conveys no information about the number of individuals that are not infected.

**Table 4.** Best-fit parameters of Equations (8) for: fixed **κ**\* = **0** (first line); all parameters adjustable (second line); fixed population of **5.0**E**4**, **5.0**E**5** and **7.5**E**5** (lines 3 to 5); fixed **q = 1** (last line).

| $u$ | $\kappa$ | $\kappa^*$ | $q$ | $\chi^2/n_{\text{dof}}$ |
| --- | --- | --- | --- | --- |
| 2430(55) | 3.0(4)E-4 | 0 | 0.56(2) | 81/42 |
| 2446(77) | 3.0(5)E-4 | $0.05 \pm 0.14$ | 0.57(3) | 81/41 |
| 5.0E4 | 8.22(6)E-5 | 4.02(3) | 0.605(7) | 138/42 |
| 5.0E5 | 1.731(1)E-5 | 8.635(7) | 0.999(2) | 194/42 |
| 7.5E5 | 1.2026(7)E-5 | 9.01(6) | 0.999(2) | 318/42 |
| 1.56837(1)E5 | 2.936(6)E-5 | 4.574(9) | 1 | 162/42 |

**Fig. 7.**
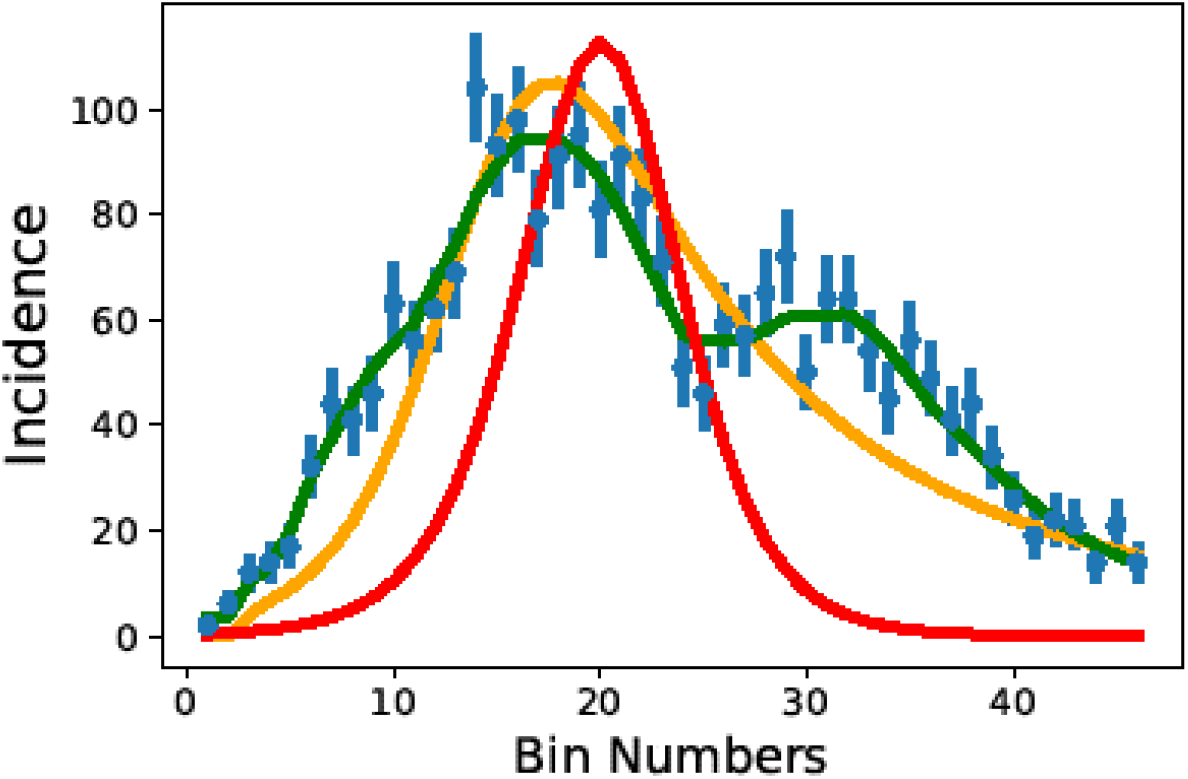
Comparison between some variations of the model. In blue are the data separated into 4-day bin and the green curve represents the fit with Equation (7). The orange curve represents the behavior of eq. 13 when q → 1. The red curve is the fit with differential Equations (8) when q = 1 and k* = 0 which corresponds to the traditional SI model.

**Fig. 8.**
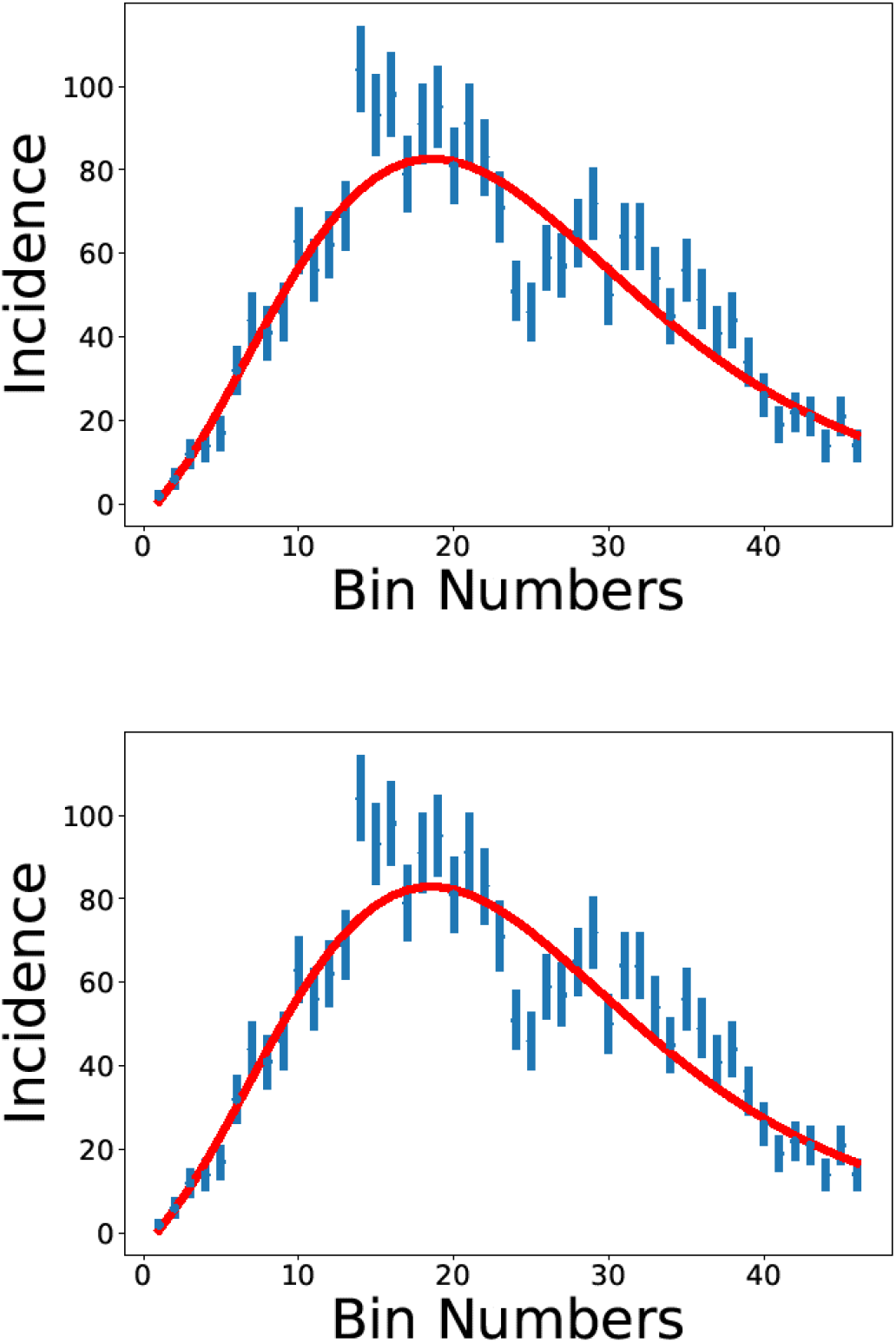
Fit for the data set separated in 4 day bins with: (top panel) k* = 0 and the other parameters left free; (bottom panel) all parameters left free.

The fitting to data can be improved, of course, by including multiple agents instead of just one agent for the entire population. The fact that *q* ≠ 1 is already an indication that the number of contacts is limited, and the homogenous mixing hypothesis of the SIR model fails.

To obtain a value for *κ*\* different from zero we need to include additional information, as an estimate of the total susceptible population, including those that will not be infected during the epidemic episode. This is not easy to do in the context of an agent-based model, but can be done if one considers the entire city as a single group and the susceptible population as the entire population in the city. This hypothesis has some degree of arbitrariness, since the city population fluctuates along the day and along the week (São Caetano is in the Metropolitan Region of São Paulo, and there is a large amount of movement between neighboring cities), therefore the meaning of the parameter *κ*\* has to be interpreted with care.

In our analysis, we study the behavior of the model for populations fixed at different values, up to those similar to the city population, estimated at *s*_*o*_ = 161,957 inhabitants (39). The results are shown in Figure (9), and additional plots can be found in the SIA. The best-fit parameters are presented in Table (4), with the results obtained showing values for *κ*\* that are not null, but that depend on the population size used. We observe that the parameter *q* also depends on the population. The fact that *q* →1 as we increase the population is due to the fact that we use one single agent even for large populations, and this result shows that the SIR is a special case of the fractal method, corresponding to the situation of a large agent. When we fix the parameter *q* = 1, the model still fits the data, but with a chi-square considerably higher than those obtained with other values of the same parameter. It is remarkable, however, that the population obtained for the best fit in this case is similar to the total population in the city. In Table (4) we observe that *κ*\* is dependent on the population size, therefore it can be determined only with the help of additional information, beyond that contained in the data on the new case time series: either fixing the population size, or determining *κ*\* by other means, as the average time in which an infected individual can transmit the virus to other individuals. In the case of COVID-19, this time is around 5 days, so we expect *κ*\* ∼ 0.2 *d*^−1^.

**Fig. 9.**
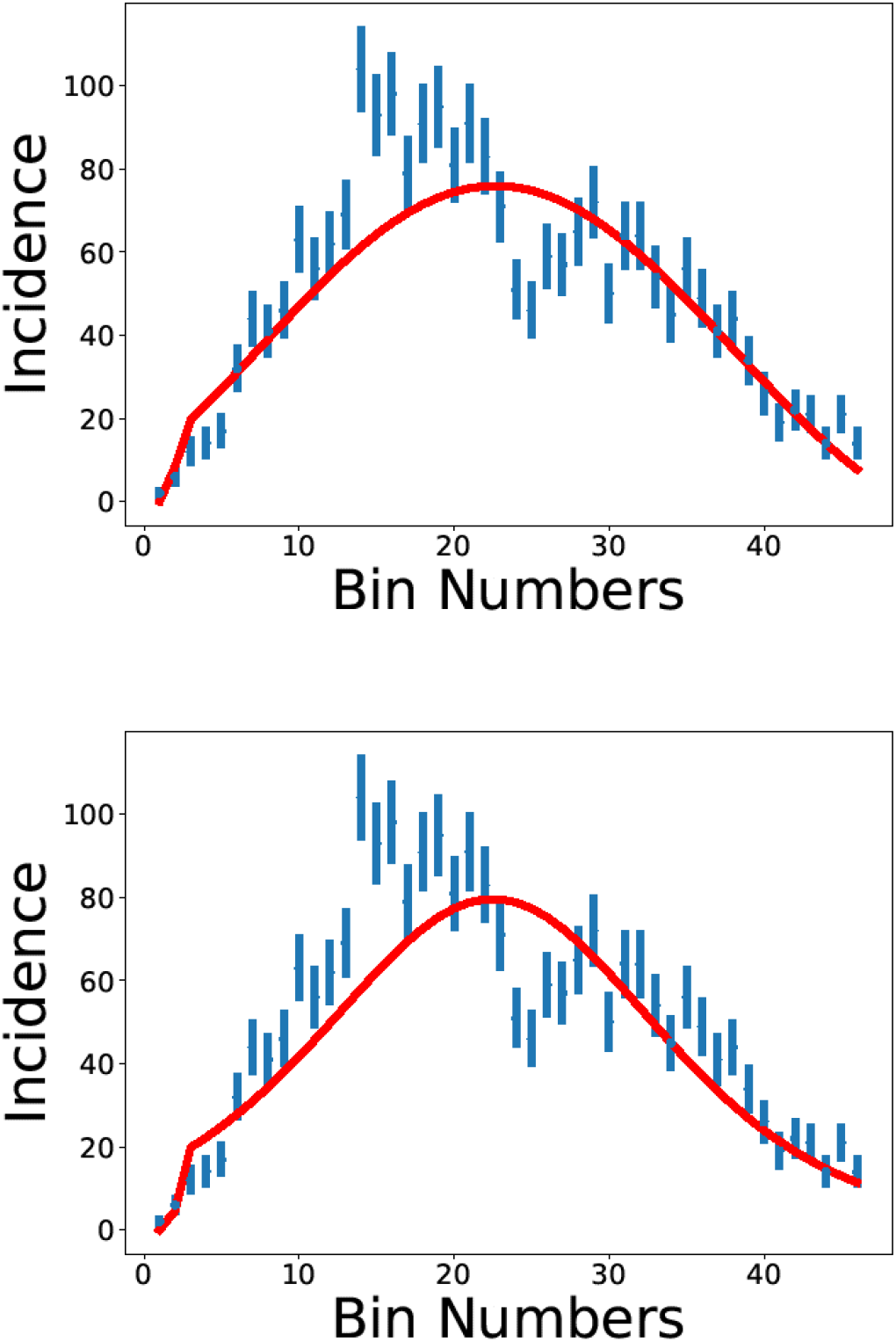
Data fit considering the data set separated in 4 day bins. In the top panel the value population is fixed to 5 .0E4. In the bottom panel we show the result for fixed q = 1 and free population.

The expected value for *κ*\* is based on the average period in which one infected individual is infecting others. Including this extra information and fixing *κ*\* = 0.2 still leads to a reasonable description of the data, as observed in Figure (10). The best-fit parameters are shown in Table (6). The result is obtained with only three agents, and when new agents are added, the fitting to the data worsens. In all cases, *q* ≠ 1, showing the importance of taking into account the limited number of contacts. Using one single agent, with *q* = 1, which reproduces the SIR model, does not result in a good reproduction of the data, as shown in Figure (10).

**Table 5.** Best fit parameters of Equation (8) obtained with bins of 4 days for orange curve and red curve of Figure (7). The first line refers to Equation (7) when (**q → 1**). The second line represents the behavior of the differential equation (8) when **q = 1** and **κ* = 0**. The parameter κ = κ_o_s^−β^, with β = 0.833 was used.

| $\kappa_0$ | $u_o$ | $q$ | $\Delta t$ | $\tau$ | $\chi^2/n_{\text{dof}}$ |
| --- | --- | --- | --- | --- | --- |
| 0.0209(5) | 2319(52) | 0.9999 | 267(1) | 0.009(2) | 196/43 |
| 8.1(2)E-5 | 1179(33) | 1 | 128(1) | 0.0103808(8) | 1091/43 |

**Table 6.** First three rows: Best-fit parameters of Equation (8) with **k* = 0. 2**, 3 agents and other parameters free. Last row: single-agent,**q = 1**.

| $u_j$ | $\kappa_j$ | $q_j$ | $R_{0j}$ | $\chi^2/n_{\text{dof}}$ |
| --- | --- | --- | --- | --- |
| 461(90) | 0.00136(40) | 0.66(9) | 3(1) |  |
| 1366(91) | 0.000377(32) | 0.74(3) | 2.5(3) | 36/33 |
| 658(62) | 0.00324(79) | 0.38(6) | 10(3) |  |
| 3555(93) | 0.000072(2) | 1 | 1.29(5) | 771/43 |

**Fig. 10.**
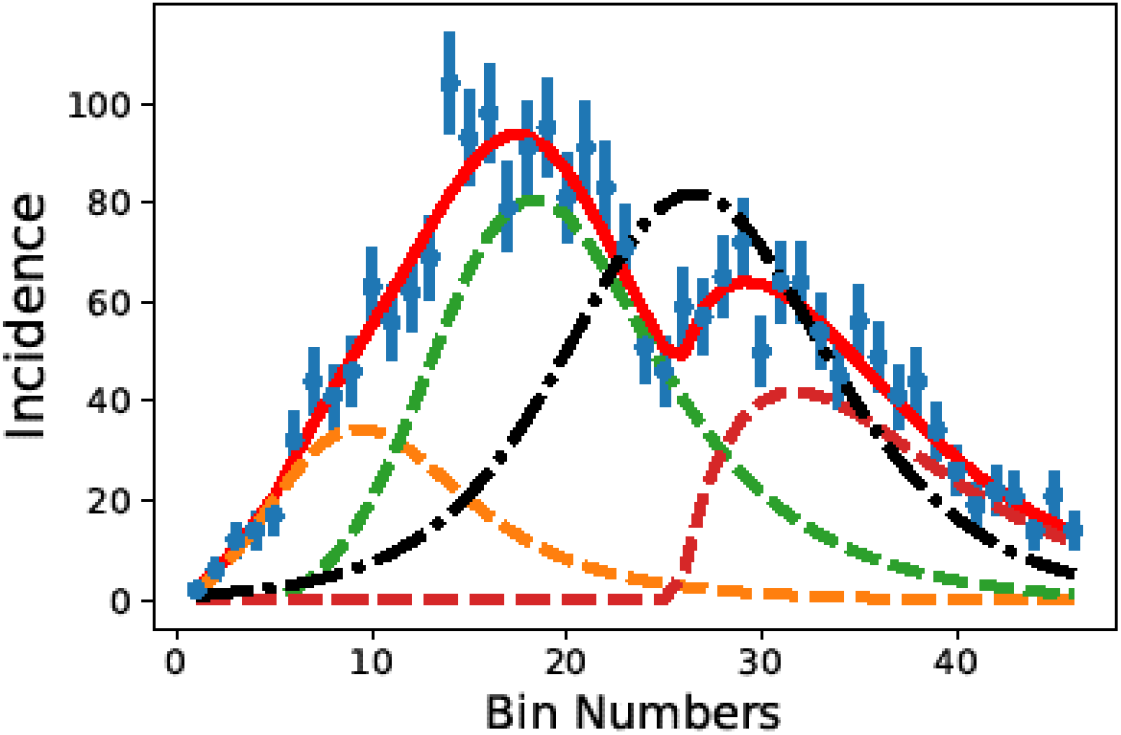
Best fit with 4-day bins setting the value k* = 0.2, 3 agents and other parameters free with background. The dashed lines represent the individual agent contributions, the continuous red line represents the total rate of infection, and the dot-dashed line represents the SIR result, with q = 1.

The best fit, with all parameters adjustable, leads to *κ*\* ∼ 0. According to Equation (8), if *κ*\* = 0 the derivative *di/dt* is proportional to *i*^*q*^, which is itself a *q*-exponential. In fact, the derivative of

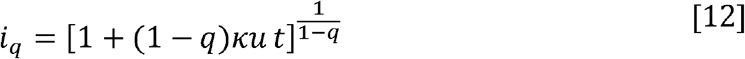

is of the form

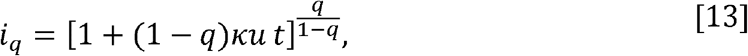

which can be written is a form similar to Equation (12), if we substitute *q* by *q′* such that

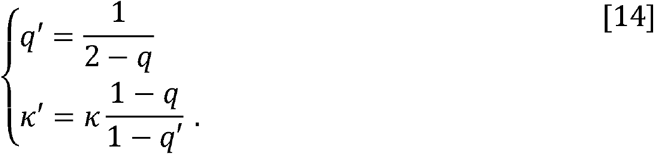

Clearly, fitting the data with a function *i*_*q*_ or with or its derivative 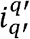 will lead to different values for the parameters *q* and *q′*, but it is possible to find a relation between the values for *q* and *κ* found in Part 1, with *q′* and *κ*′ found in Part 4, using *κ*\* = 0. The fact that the q-exponential function gives good fits to the data is an indication that the mechanism derived for the small group can, indeed, be applied to large groups. This is the fundamental hypothesis adopted in the fractal dynamic model, so the results obtained here corroborate the fractal assumption.

## Conclusion

In this study we used original data on the number of newly infected individuals in the city of São Caetano do Sul, in the metropolitan area of São Paulo. The data reports the number of new cases on a daily basis and contains the dates of initial symptoms and the geographical localization of an individual’s residence.

We found that, to accurately describe the observed epidemic in this city, we need to invoke a mechanism that limits the number of contacts between infectious people and others in the population; in other words, the supposition made in conventional SIR and SI models that people mix randomly across whole cities gives an unsatisfactory explanation of COVID-19 dynamics in São Caetano do Sul. We suggest that the same may be true of other cities in Brazil and perhaps more widely.

The contact process investigated here produces a fractal model with scale-free dynamics., The model has the possibility of including new agents (i.e., groups of close contacts) in the transmission process, while keeping the total number of parameters small enough to obtain information on the epidemic from the available data.

With these data, we checked the fractal dynamic model by various approaches. We found that the fractal model gives the expected power-law distributions of variables as a function of the population size, as obtained for the infection rate, *κ* and for the period of infection, *Δt*. By incorporating these power-laws in the model, we can investigate the effects of social distancing by analysing parameter *q*, which is determined by the average number of contacts made in practice. Our results show that, in the first wave of the epidemic, the average number of contacts was approximately 2, while in the second wave it increased to approximately 5. This result, obtained by using the model, and based solely on the time distribution of the newly infected individuals, is corroborated by an independent analysis of social-distancing, based on the movement of individuals in the city. For each period, there is a variation of the number of contacts for each agent, with super-spreader agents being observed in the second wave, as can be observed in Figure (4), with a number of contacts larger than 10, corresponding to values of *q* > 0.9 (see also Table 3).

The data on geographic localization of each individual’s residence allowed us to check the main conceptual aspect of the model, namely transmission among groups of people in close contact. The assumptions of the model are in agreement with the observed geographical clustering of cases. The analysis of the least distance among successive cases indicates that the average distance between newly infected individuals and those previously infected is approximately 200 meters. The clustering model allows us to identify the small groups of contagion in the city, and verify that the location of the clusters coincides with places of intense flux of people. Our model therefore gives a realistic description of the spread of infection at small as well as large geographical scales.

The comparison of the fractal dynamics model with the SIR and SI models was made using the fact that, as *q* → 1, the fractal differential equations reduce to those of the SIR model (with *κ*\* ≠ 0) and SI model (with *κ*\* = 0). The best description of the data, obtained in Part 4 of our analysis when all the parameters of Equation (8) are free to adjust to the data, results in *q* ∼ 0.5 and *κ*\* = 0. This gives an estimated number of close contacts of ∼2, as noted above. This result is an indication that the distribution of new cases is best described by a q-exponential function, and thus the best solution found in Parts 1 or 2 and in Part 4 are equivalent.

We conclude that new cases are usually generated at small distances from previous cases, and that the spread of COVID-19 is based on small and highly localized clusters of infection. The use of networks to simulate the dynamical evolution of an epidemic process allows a more detailed description of the process, and confirms that those tools are useful to analyse socioeconomic aspects of modern life, as already observed in other studies (40–43), in particular, scale-free or fractal networks, have been identified in several domains (34, 35, 44–46). For COVID-19 control, the practical value of our model is in help to choose, for example, between societal lockdown versus improved social distancing, and to optimize the size of the population affected by those decisions and the time necessary for such measures to be effective.

X’As an hypothetical example, consider the problem of deciding to lockdown an entire region to mitigate the epidemic process. One of the main parameters to be determined is the duration of the lockdown, and the model described here can bee used to determine the optimal duration of a lockdown in any given geographical region Let us suppose that, in that region, the average household is formed by four individuals. According to our findings, the infectious period 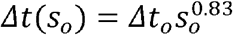, where *s*_*o*_ is the average household size. According to the power-law behaviour obtained in our analysis, the infectious period in an agent of size *s*_*o*_ is given by 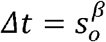, with *β* = 0.833. If *t*_*o*_ is the time during which one individual can infect others, for a group with *s*_*o*_ = 4 we have *Δt* ∼ 3*Δt*_*o*_. If the period in which one individual can infect others is, say, 10 days, then the optimal duration of lockdown is approximately 30 days.

In conclusion, we have shown that a fractal dynamic model for the COVID-19 infection process gives results consistent with epidemic data for the city of São Caetano do Sul, and at different scales of population size. An additional, scale-free feature of the model, the independence of the average number of contacts and population size, allows us to evaluate the effects of social distancing, based only on the time series of newly infected individuals. The possibility of separating the transmission probability (a property of the virus), from the average number of contacts made by infectious individuals (a property of the human population) is a benefit of the fractal dynamic model, which is shared by other agent-based models of social behaviour.

## Data Availability

All data used in the work is available in the manuscript and supplementary material.

## Appendix

In this section, we describe the methods used to collect the data and for the analysis. The latter is divided into four parts: a test on the model consistency; an investigation on the properties of the spreading dynamics; a cross-check of the results of our analysis with additional information; and a comparison with the SIR model.

### A. The data

The data were collected in São Caetano do Sul, a city in the metropolitan region of São Paulo, Brazil, with an estimated population of 161,957 inhabitants (39) and one of the highest Human Development Index values in the country. In April 2020, a primary care initiative offering COVID-19 care to all residents was created. Residents of the municipality aged 12 years and older with suspected COVID-19 symptoms were encouraged to contact the dedicated Corona São Caetano platform via a website (access at \url{https://coronasaocaetano.org/}) or by phone. They completed an initial screening questionnaire that included clinical and socio-demographic data. Patients meeting the suspected COVID-19 case definition were further evaluated by a health care professional and offered a home visit for sample collection for PCR testing (48). The São Caetano platform routinely collects the patient addresses and transforms them into latitude and longitude values using an API from the Here company (Chicago, USA). For this study we received the lat/long values with a decreased precision of 2 decimal digits that corresponds to an area of 1.2km^2^. We also received the exact date of symptom initiation for each patient.

The study was approved by the local ethics committee (CAAE 32424720.8.0000.0068). The committee waived the need for informed consent and allowed the development of an analytical dataset with no personal identification for analysis.

#### A.1. Social Distancing

Social distancing adherence was investigated in a validation step of this study, and considered two different datasets. Both datasets provide a relative measure of the number of people that stayed home during a reference day, resulting in what we call the Social Isolation Index. The government of São Paulo issues an official calculation of the index based on radio signals of mobiles from network stations of the four main providers in Brazil (Oi, Vivo, Claro, Tim). This leads to an estimate of the user’s location within the order of meters, depending on the number of nearby base stations. The home place is decided based on the location the user has spent the night (from 22h to 2h), and movements outside of the 200m range from the house are triggered as a breach of isolation. This dataset is publicly available on a city level at the official government website (https://www.saopaulo.sp.gov.br/coronavirus/isolamento/). The second dataset uses geolocation data collected from a technology company called InLoco, which in 2020 collected positions of around one fourth of the Brazilian population. Home location is then decided in a similar way as for the network providers data, but the breach in isolation is decided according to a change in position of around 460m, which explains the higher percentages of isolation verified in Figure (5). It is provided in the same way as the government data, as a Social Isolation Index aggregated by cities (the dataset is included in the supplementary material for this paper). During this pandemic, several studies have been using this dataset as it provides a good metric of social distancing (37, 50–53).

#### B. The analysis

**Step 1:** Initially, we group the number of infected individuals in bins of increasing width, starting from bins 4-day large, until the reduced *χ*^2^ reaches a minimum value when described by a single agent. We found that it happens for bins 30-day large.

The fitting gives the parameters of Equation (7) *u*_*o*_, *κ* and *t*_1_. The next step is to divide the bin width by two, double the number of agents, and repeat the process of fitting, getting, as a result, the parameters for each of the two agents, as well as the global parameter, *c* for this bin width. The process is repeated, doubling the number of agents and fitting the parameters until a good fit is obtained with a bin width of four days. At the end of this part, we have fitted in total 12 agents and obtained 36 best-fit parameters for those agents. The analysis of the behaviour of the parameters *κ* gives a power-law distribution, as we show in the next section.

The details can be seen in the supplementary material online.

**Step 2:** In this step we adopt, in Equation (7), the power-law behaviour of *κ*(*u*_*oj*_), that is, *κ*(*u*_*oj*_) = *κ*_*o*_*u*^*β*^, with the value of *β* found in Step 1. The parameter *κ*_*o*_ is global, being the same for all agents with a given scale. However, the number of free parameters per agent in this step remains the same because now we include *q* as a free parameter in the fitting process. With this, we can infer from the data the average number of close contacts that have those individuals who are transmitting the virus.

The procedure adopted here is similar to that of Step 1, except that now, for each peak, the free parameters in the fitting procedure are *u*_*oj*_, *t*_*j*_ and *q*_*j*_, while *κ*_*o*_ and *c* are global parameters, being the same for all agents for the same bin width. This analysis allows us to observe how the value of *q* changes from one group to the other and as the epidemic process evolves. The results can be seen in the supplementary material online.

**Step 3:** In this part we perform further analyses of the data, aiming to verify the robustness of the fractal model used in this work. One of the assumptions in our model, and its main difference with respect to the SIR model, is that transmission occurs only among individuals in the groups of close contact, no matter the scale of the agent.

A consequence of this assumption is that the transmission of the virus may present a strong correlation with the geographical region where each infected agent lives. As the data we use have the location of residence of the infected individuals, we can investigate the correlation between time of infection and localization.

A sequence of 2D plots indicates, by using different symbols, the groups of individuals already infected and the new infected at each day. To make a clear visualization, we just investigate the initial period of the infection in São Caetano do Sul. We calculate the least distance between newly infected and infected individuals in the city at different periods of time for the first wave of contagion. The histograms of least distance evidence the spatial reach of the spreading dynamics.

Finally, we created a method to identify clusters of infection among the infection cases. The clustering method uses time and space correlations to identify the sequence of contagion in the different agents. The method consists in associating to the geographical localization the time elapsed between successive cases of infection, by a clustering method.

Consider that at day *t* we have a set of clusters of infections, *Ct* .To each cluster we associate a vector position 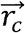 given by the average position of the individuals in that cluster. For each newly infected individual in day $t$ we attribute a probability to belong to a cluster in the set given by

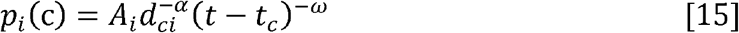

where *α* and *ω* are parameters of the clustering method that are adjusted to reproduce the least-distance distributions and the time of contagion for the different agents. Here we use *α* = 3.5 and *ω* = 2. *A*_*i*_ is a normalization constant to ensure that the probabilities add up to one.

We also define a threshold value, *L*, for the acceptance in a cluster. If all values *p*_*i*_(*c*) < *L*, then the individual forms a new cluster, which is included in the set *C*_*t*+1_ with all other clusters already in the set *C*_*t*_. If there is one or more values *p*_*i*_(*c*) > *L*, with *c is in Ct*, then the individual is included in the cluster with the highest probability. When a new individual is included in a cluster *c is in Ct*, the cluster position is recalculated as the average of the positions of all individuals in that cluster, and the time *tc* is set to *t*.

**Step 4:** The comparison of the fractal model and the SIR and SI models is done by adopting Equations (8),and fitting the data on new cases by fitting -*du*_*j*_*/dt* to the series of new cases. This is an important difference with respect to the procedure used in Part 1 and Part 2, and is demanded by the necessity of using the SIR and SI methods of analysis. In this case, instead of fitting the q-exponential function to the data, its derivative is used in the fitting process. Since the derivative of a q-exponential is itself a q-exponential, this will just result in a slight different value for *q* and *κ*. In this part of the analysis, we investigate the role played by the free parameter, *κ*^*^ its relation with the population size, and how all these parameters depend on the value for *q*. We use the chi-square as a measure of the quality of the fitting and to discuss about what of the models best describe the data. Since SI and SIR are not agent-based models, we conduct the analysis using one single agent to describe the entire population.

## ACKNOWLEDGMENTS

A D is partially supported by the Conselho Nacional de Desenvolvimento Científico e Tecnológico (CNPq-Brazil), grant 304244/2018-0, by Project INCT-FNA Proc. No. 464 898/2014-5 and by FAPESP grant 2016/17612-7. PSP is supported by FAPESP grant 16/18445-7. V H N is supported by CNPq, grant 304714/2018-6. This work was supported by a Medical Research Council-São Paulo Research Foundation (FAPESP) CADDE partnership award (MR/S0195/1 and FAPESP 18/14389-0) (https://caddecentre.org). This work was supported by a Medical Research Council-São Paulo Research Foundation (FAPESP) CADDE partnership award (MR/S0195/1 and FAPESP 18/14389-0) (https://caddecentre.org); Wellcome Trust and Royal Society (N.R.F.: Sir Henry Dale Fellowship: 204311/Z/16/Z).

