## Supplementary material for "Scale-free dynamics of Covid-19 in a Brazilian city": PNAS-SIA.pdf

September 10, 2021

#### Mathematical description of the fractal spreading model

An epidemiological model to describe the spreading dynamics of COVID-19 was proposed in [1], where the fractal aspects of the virus transmission process observed in the analysis of real data on the transmission of the disease were included. Rather than an exponential increase, as would be expected by the SIR model, a q-exponential behaviour is obtained, which means that the spreading is not as fast as one could predict by using standard SIR models, and is in fact similar to the modified model proposed in [2]. The non-exponential behaviour is important to be considered in evaluations of the impact of different policies to combat the epidemic spreading on the rate of contamination. Roughly speaking, the fractal approach assumes the scale invariance of some complex structures of the epidemic dynamics, dividing the infected population into different groups at different scales. With such an assumption, the dynamics of the epidemic spreading over a large population can be related to the process in a small group. The number of parameters necessary to describe the complex dynamics is reduced.

The function that describes the number of new cases in the fractal model can be written as

$$\frac{di}{du}(\tau,N;u) = \left(1 + \frac{\tau u}{N}\right)^{N-1} \tag{1}$$

where $u$ is the susceptible population, $\tau$ (assumed small) is the transmission probability and $N$ is the average number of close contacts of the individuals in that population. To extend the spreading mechanisms to larger populations, we assume that the population is formed by a set of agents, identified by the index $j$, with approximately the same size, $N$. Following Ref. [1], we adopt the parameter $q$ such that $1 - q = N^{-1}$, which leads to the differential equations of the fractal model, given by (for $t > t_{oj}$)

$$\left\{\begin{aligned} \frac{di_j(t)}{dt} &= \kappa_j i_j^q(t)\, u_j(t) + \kappa_j i_j^q(t)(t-t_{oj})\dot{u}_j(t) \\ \frac{du_j(t)}{dt} &= -\kappa_j i_j^q(t)\, u_j(t) \\ \frac{dr_j(t)}{dt} &= -\kappa_j i_j^q(t)(t-t_{oj})\dot{u}(t) \end{aligned}\right. , \tag{2}$$

where $i_o$ is the initial number of infected agents, which we assume to be $i_o = 1$. We assume that the susceptible population of the agent $j$ is given by $u_j(0) = u_{oj}$, to be determined in the data analysis, and that the rate of transmission, $\kappa_j$, may vary from one agent to the other. The time of introduction of the virus in the agent $j$ is $t_{oj}$, corresponding to the instant when the first individual of the group is infected. The removed population $r_j$ corresponds to the individuals that are placed out of the spreading dynamics because they can no longer transmit the virus either because they have recovered or died, or because they no longer have susceptible individuals in their neighborhood.

The expression for $i(t)$ is known in the non-extensive statistics proposed by Tsallis [3] as the q-exponential distribution. In this statistics, mean values are determined by expressions as

$$< a >= \int a p^q(a) da \tag{3}$$

where $q$ is called entropic index, and plays a role similar to the parameter $q$ in the fractal model , and $p(a)$ is the probability density. . The fact that we have q-exponential distributions and that the probability to find an infected individual appears in powers of $q$, shows the close connections between the Tsallis statistics and the spreading dynamics in a fractal structure.

Aside from the evident agent-based approach of the model described by the equations above, a few additional remarks are necessary. The number of parameters used for each agent is large, with $q$, $\kappa_j$, $u_{oj}$ and $t_{oj}$ being undetermined. The parameter $N$, related to the agent size, can be evaluated by other means as a general property of the network, or can be considered as a quantity that may vary from one agent to the other. In any case we are left with a large number of free parameters that increases linearly with the number of agents.

Observe that the term corresponding to the rate with which the removed population increases, that appears in the first and in the third equations, can be written as

$$\frac{dr_j(t)}{dt} = -[\kappa_j i_j^q(t) u_j(t)]\kappa_j(t-t_{oj}) i_j^q(t)\,. \tag{4}$$

when the second equation is used. This term is very different from its correspondent in the SIR model. Here, the removed population is determined by the epidemic process inside each agent, and takes into account the fact that after some time of the first infection, all individuals in the group will be already infected through one of the possible modes of infection. Another aspect that differentiates the fractal model from the usual SIR model is the explicit time dependence of the epidemics, introduced by the used of the term $\tau_j = \kappa_j(t - t_{oj})$. Within the fractal model, the instant when the virus is transmitted to other individuals in the agent is aleatory.

We can simplify the expression above by assuming an average contamination rate for the term between brackets, multiplied by $\kappa_j(t-t_{oj})$. Taking the average of this term we have

$$\kappa_j(t - t_{oj})\kappa_j i_j^q(t)u_j(t) = -\kappa_j(t - t_{oj})\dot{u}_j(t) \sim -\langle \kappa_j(t - t_{oj})\dot{u}_j \rangle \tag{5}$$

which corresponds to a linear approximation (because $\dot{u}(t)$ is substituted by a constant) of the behaviour of the susceptible population. Considering the period during which the virus is being transmitted inside the agent, $\Delta t_j = t_{fj} - t_{oj}$, with $t_{fj}$ being the time when the new cases of infection in the agent cease, we have, for the first two terms at the right-hand side,

$$\langle \kappa_j(t - t_{oj}) \rangle = \langle \kappa_j \Delta t_j \rangle \frac{\langle t - t_{oj} \rangle}{\Delta t_j} \,. \tag{6}$$

But $\kappa_j \Delta t_j = \tau$ is constant, and $\langle t - t_{oj} \rangle \sim \Delta t_j/2$, since the distribution is just slightly asymmetric. Therefore, with the approximations above, we obtain

$$\frac{dr_j(t)}{dt} = \kappa_j^\dagger i_j^q(t) \,. \tag{7}$$

where

$$\kappa_j^\dagger = \frac{\tau}{2} |\langle \dot{u}_j \rangle| \,, \tag{8}$$

is completely determined by the parameters of the epidemic dynamics, with $\tau = \kappa_j \, \Delta t_j$, where $\Delta t_j$ is the time during which the virus is circulating in the agent $j$.

The considerations made above show that the fractal model has a structure that is similar, in some aspects, with the SIR model. However, contrary to the SIR model, in the fractal model the parameter of the removed population, $\kappa^\dagger$, is completely determined by the other parameters in the fractal dynamics. This happens because we are considering that the total population, $u_o$, will be infected, which is an unrealistic situation since, in practice, it is observed that part of the susceptible population will not be infected in the period the virus is circulating in the group. We can improve the fractal model in this aspect by including a new parameter that refers to those individuals that will be removed from the infected population before all the individuals will be infected. Defining

$$\kappa_j^* = (1 + \gamma)\kappa^\dagger \tag{9}$$

we introduce the new parameter $\gamma \geq 0$ that corresponds to the removed population, which allow us to obtain a fractal model similar to the SIR model when we substitute $\kappa^\dagger$ by $\kappa^*$ in Equation (7). Accordingly, the infected population equation will be

$$\frac{di_j(t)}{dt} = \kappa_j i_j^q(t - t_{oj})\, u(t - t_{oj}) - (1+\gamma)\kappa_{qj}^\dagger i_j^q(t - t_{oj}) \; . \tag{10}$$

Observe that the equation above can be obtained by considering that the susceptible population in the group is $s_j(t) \geq u_j(t)$ in the expression for $i(t)$. If $\gamma > 0$, the susceptible population will be larger than the total infected population, as it usually happens in the SIR model. The set of equations that describes the fractal model equivalent to SIR model is

$$\begin{cases} \dfrac{di_j(t)}{dt} = \kappa_j i_j^q\, s_j - \kappa_{qj}^* i_j^q \\ \\ \dfrac{ds_j(t)}{dt} = -\kappa_j i_j^q\, s_j \\ \\ \dfrac{dr_j(t)}{dt} = \kappa_{qj}^* i_j^q \,, \end{cases} \quad . \tag{11}$$

for $t \geq t_{oj}$. For $q = 1$, the equations above reduce to the SIR model equations. In this case, the usual notation is obtained by using the parameter $\beta_j$ such that $\kappa_j = \beta_j / s_{oj}$.

From the Equations 11, it is possible to understand another difference between the fractal model and the SIR model. While in the latter the number of close contacts increases as the population size increases, in the fractal model the two numbers are uncorrelated. This means that the number of close contacts of an individual does not increase as the population increases, reflecting the fact that the number of close contacts that an average person has does not vary significantly if that person is in a metropolis or a small city. This model describes many features and details of the COVID-19 dynamics [1].

While the transmission probability depends on the features exhibited by the different virus variants and is mostly out of our control, the number of close contacts can be reduced by social distancing and has an effective impact on the number of new cases. Therefore social distancing will be effective to reduce the number of new cases even for the new variants of SARS-COV-2 with higher transmission probability.

It is important to stress that a clear separation between transmission probability and social distancing is never possible. For instance, the use of protective devices, such as masks, is to be considered as a reduction in the transmission probability or as social distancing? This is a question that is not even posed in the SIR model, as it does not allow a separation between the two contributions. The fractal model, however, does allow such a separation of the the two contributions, as we will show below. Therefore, the question above becomes relevant and has to be addressed in some way. Our approach, in this work, is to consider as transmission probability only those features that are intrinsically related to the virus characteristics, and attribute to social distancing all features of the spreading dynamics that can be controlled by human action. Of course, this definition is arbitrary, and still may not solve completely the problem of separating transmission probability from social distancing. An example is the vaccination process, which represents a human action that interferes in the virus capability to infect individuals. However, to the present work, this aspect is irrelevant, since no vaccination took place for the entire period of analysis.

#### Calculation of the distance between new cases and clusters

This analysis is performed by making 2-dimensional plots of the residence location of the infected individual along time. The residence location is obtained form the longitude, $Lo$, and latitude, $La$ (in radians), of residence by defining the vector

$$\vec{r}_i(x,y) = R_T Lo\,\hat{\imath} + R_T La \cos(Lo)\,\hat{\jmath}\,, \tag{12}$$

with $\hat{\imath}$ indicating the North-South direction and $\hat{\jmath}$ the East-West and $R_T$ is the Earth radius at the Equator line. The sum of all cases identified up to the day $t$ are collected in a set $I_t$, where $t$ is the day when the individual $m$ reported the first symptoms. The distance to the previous cases is calculated by

$$d_{mn} = \sqrt{(\vec{r}_m - \vec{r}_n)^2}\,, \tag{13}$$

where $n$ indicates each of the individuals in the set $I_t$. The least distance, $d_m$, is the minimum value in the set of distances for the new case, $c$, that is,

$$d_m = \min_{n \in I_t}\{d_{mn}\}. \tag{14}$$

### Additional plots and graphs

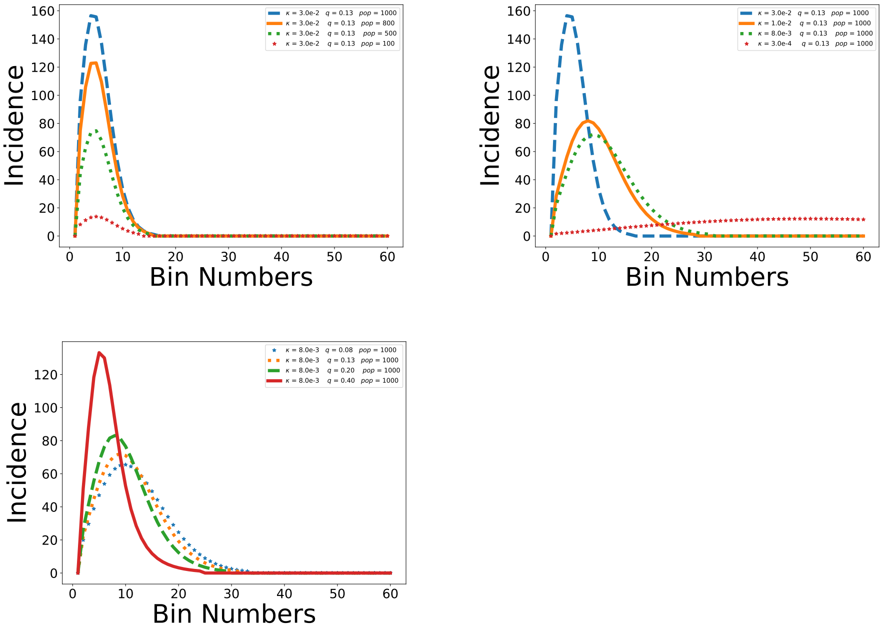


Figure 1: Epidemic evolution according to the fractal dynamics model using the equation 7. In the top-left panel, the population size varies, with all other parameters constant, and in the top-right panel, the transmission probability varies. At the bottom, the dependence of the model with the parameter $q$.

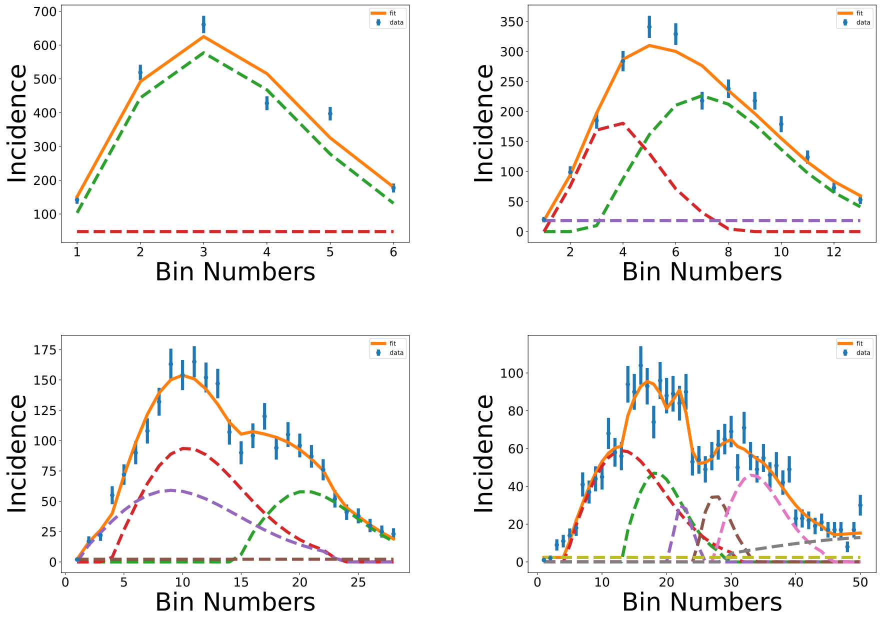


Figure 2: The results of the fitting procedure for (top-left) 30-day bin, (top-right) 15-day bin, (bottom-left) 7-day bin and (bottom-right) 4-day bin analysis. The total number of infected and the contributions of each agent are shown, as well as the background contribution. The best parameters are shown in Table 1 in the main text.

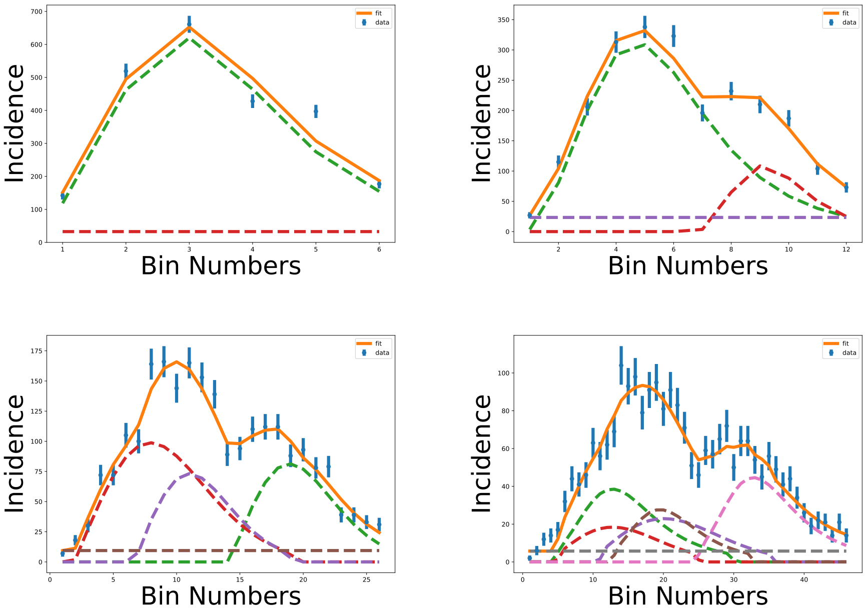


Figure 3: The results of the fitting procedure for (top-left) 30-day bin, (top-right) 15-day bin, (bottom-left) 7-day bin and (bottom-right) 4-day bin analysis. The total number of infected and the contributions of each agent are shown, as well as the background contribution. Here $\kappa$ follows the power-law $\kappa = \kappa_o u^{\beta}$ with $\beta = -0.833$. The best parameters are shown in Table 2 of the main text.

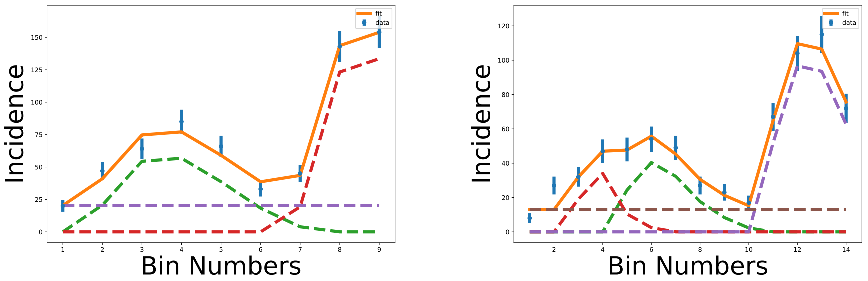


Figure 4: The results of the fitting procedure for (left) 7-day bin and (right) 4-day bin analysis for second wave. The total number of infected and the contributions of each agent are shown, as well as the background contribution. Here $\kappa$ follows the power-law $\kappa = \kappa_o u^{\beta}$ with $\beta = -0.833$.

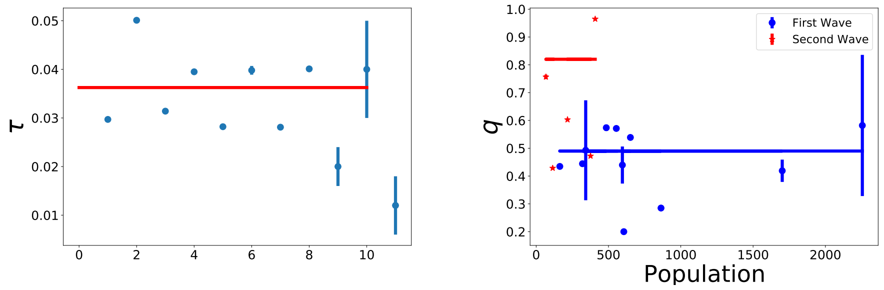


Figure 5: Distribution of the $\tau = \kappa\Delta t$ parameter for the first wave with $\langle\tau\rangle = 0.03625(7)$ (left). Best values of $q$ as a function of agent size (right).

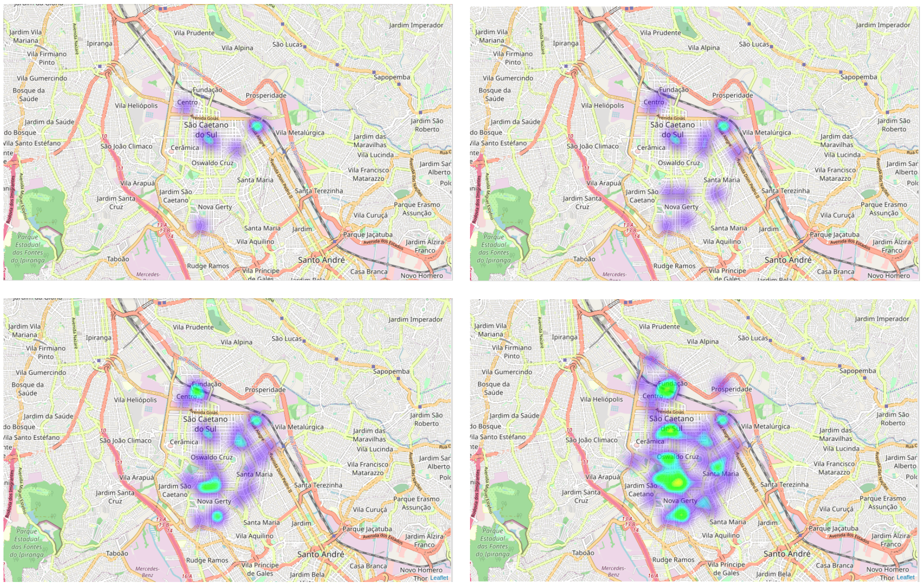


Figure 6: The evolution of the virus in the city of São Caetano do Sul through a heat map. The images show four moments after the first infection, starting at 7 days (top-left), 15 days (top-right), 30 days (bottom-left) and 60 days (bottom-right).

Table 1: Best fit values for the parameters in Second Wave obtained with bins of 4 and 7 days. In these cases the $q$ and $\kappa_0$ parameter is free and $\kappa$ follows the power-law $\kappa = \kappa_o u^\beta$ with $\beta = -0.833$ as shown in Figure 4. The parameter $c$ is a constant that represents the number of individuals that were infected out of the city limits. The Contamination period ($\Delta t$) is the number of days between the first contaminated of the group until 95% of the group is contaminated and $\tau = \kappa\Delta t$.

| $\kappa_0$ | $u$ | $q$ | $\Delta t$ | $\tau$ | $\chi^2/n_{\rm dof}$ |
|---|---|---|---|---|---|
| 4-day bin | 114(11) | 0.429(4) | 15(1) | 0.14(1) | |
| $\kappa_0$ = 0.0.48071(15) | 67(1) | 0.756(8) | 9(1) | 0.1370(7) | 14/3 |
| $c = 13 \pm 1$ | 376(1) | 0.472(1) | 24(1) | 0.0819(2) | |
| 7-day bin | 216(2) | 0.602(1) | 33(1) | 0.0645(9) | |
| $\kappa_0$ = 0.172520(13) | 408(1) | 0.9647(7) | 34(1) | 0.03922(4) | 6/1 |
| $c = 23 \pm 1$ | | | | | |
